# COVID-19 Recurrent Varies with Different Combinatorial Medical Treatments Determined by Machine Learning Approaches

**DOI:** 10.1101/2020.07.29.20164699

**Authors:** Jia Huang, Song Zhai, Fangfan Ye, Song Wang, Manfei Zeng, George Way, Vipul Madahar, Tengfei Zhu, Liping Qiu, Zehui Xu, Manhua Ye, Lei Liu, Xinping Cui, Jiayu Liao

## Abstract

Various medical treatments for COVID-19 are attempted. After patients are discharged, SARS-CoV-2 recurring cases are reported and the recurrence could profoundly impact patient healthcare and social economics. To date, no data on the effects of medical treatments on recurrence has been published. We analyzed the treatment data of combinations of ten different drugs for the recurring cases in a single medical center, Shenzhen, China. A total of 417 patients were considered and 414 of them were included in this study (3 deaths) with mild-to-critical COVID-19. Patients were treated by 10 different drug combinations and followed up for recurrence for 28 days quarantine after being discharged from the medical center between February and May, 2020. We applied the Synthetic Minority Oversampling Technique (SMOTE) to overcome the rare recurring events in certain age groups and performed Virtual Twins (VT) analysis facilitated by random forest regression for medical treatment-recurrence classification. Among those drug combinations, Methylprednisolone/Interferon/Lopinavir/Ritonavir/Arbidol led to the lowest recurring rate (0.133) as compared to the average recurring rate (0.203). For the younger group (age 20-27) or the older group (age 60-70), the optimal drug combinations are different, but the above combination is still the second best. For obese patients, the combination of Ribavirin/Interferon/Lopinavir/Ritonavir/Arbidol led to the lowest recurring rate for age group of 20-50, whereas the combination of Interferon/Lopinavir/Ritonavir/Arbidol led to lowest recurring rate for age group of 50-70. The insights into combinatorial therapy we provided here shed lights on the use of a combination of (biological and chemical) anti-virus therapy and/or anti-cytokine storm as a potentially effective therapeutic treatment for COVID-19.

## Introduction

Since the outbreak of COVID-19, it has quickly spread to more than 200 countries worldwide as an unprecedented global pandemic. Up to now, the global number of confirmed patients has risen to more than 10 million, and the death rate has reached 510,632 (death rate ∼ 4.89%)(1). While many vaccines are still under development with very encouraging preliminary results, no specific anti-SARS-CoV-2 drug has proved to be effective except Remdesivir for an emergency use authorized by FDA(2, 3). Although Remdesvir shows treatment benefits by reducing hospitalization time for 31%, the reduction of the death rate has not reached statistical significance (8.0% vs.11.6%, 0.059)(4). Moreover, the SARS-CoV-2 recurrence has significant impacts on disease management and society quarantine policy after patients are clinically cured and discharged. The report of the 116 recurring cases identified through the NP swabs in South Korean caused serious concerns about reactivation or reinfection of SARS-CoV-2(5). In another report, one hospitalized patient and 3 medical personnel were tested positive again after medical treatments and fulfilling discharge clinical criteria and being followed up for a period of quarantine(6). One patient, who was ready-for-discharge after three consecutive negative NP swabs, died from cardiac arrest with SARS-CoV-2 viruses remaining in pneumocytes(7). In addition, the asymptomatic transmission of SARS-CoV-2 from asymptomatic carriers with normal chest computed tomography (CT) or CT-positive patients exacerbate the concerns of COVID-19 recurrence(8). To date, no study on medical treatments affecting SARS-CoV-2 virus recurrence has been published in the scientific literature. Here, we report the clinical, radiological, laboratory, and drug treatment findings of 93 recurring patients from 414 patients in Shenzhen, along with our machine learning approaches for identifying the best drug combinations that reduce recurring rates in all population, different age groups and obese patients.

## Results

### Patients

We collected data from The Second Affiliated Hospital of Southern University of Science and Technology, with COVID-19 patients who were admitted between January 1, 2020 and February 16, 2020, and completed their hospital course (discharged alive) by March 26, 2020 and followed up by May 5, 2020. Three patients who died during the hospitalization were excluded from the analysis. This cohort included 414 patients. The mean (SD) age was 45 (17.7) years, 47.1% of the patients were men. 74.6% of patients had moderate disease, 3.9%, 17.6% and 3.9% had mild, severe and critical disease, respectively. Ground-glass opacity and pulmonary infiltration were the most common imaging characteristics (Table 1). Medical therapy included Interferon (82.4% of the patients), Lopinavir/Ritonavir (77.3%), Arbidol (27.3%), Ribavirin (19.3%), Oseltamivir (14.7%), Favipiravir (8.5%), and Hydroxychloroquine (6.3%). Methylsprednisolone was used in 23.9% of the patients, and Tocilizumab was used in 2.2% (Supplementary Table 1). At the follow-up, 93 (22.5%) recovered patients were found to be positive with NP swab RT-PCR tests after a median of 21 days recurring interval. Among them, 19.4% and 3.2% had experienced second and third recurrences after a median of 9 and 8 days intervals, respectively. After discharge, the median lengths of virus shedding at the 1st, 2nd, and 3rd recurrence were 7, 5, and 7 days, respectively (Figure 1A,B). Cough (16%) and sputum (11%) were the main symptoms of recurring patients at follow-up. Minority of them had abnormal biochemical parameters. The lesions (83.8% of recurring patients) were gradually absorbed in chest imaging (Supplementary Table 2).

**Table 1:**
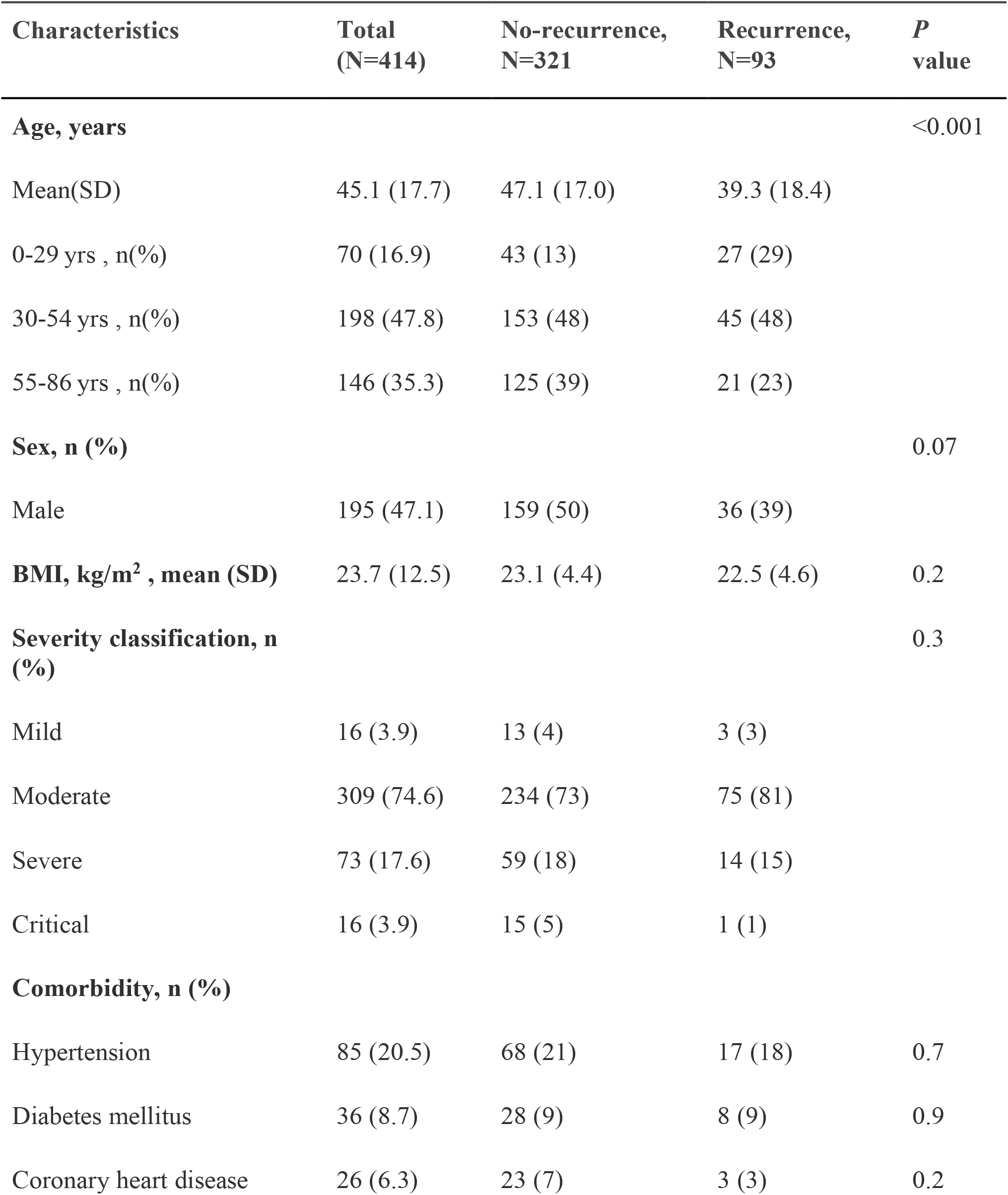
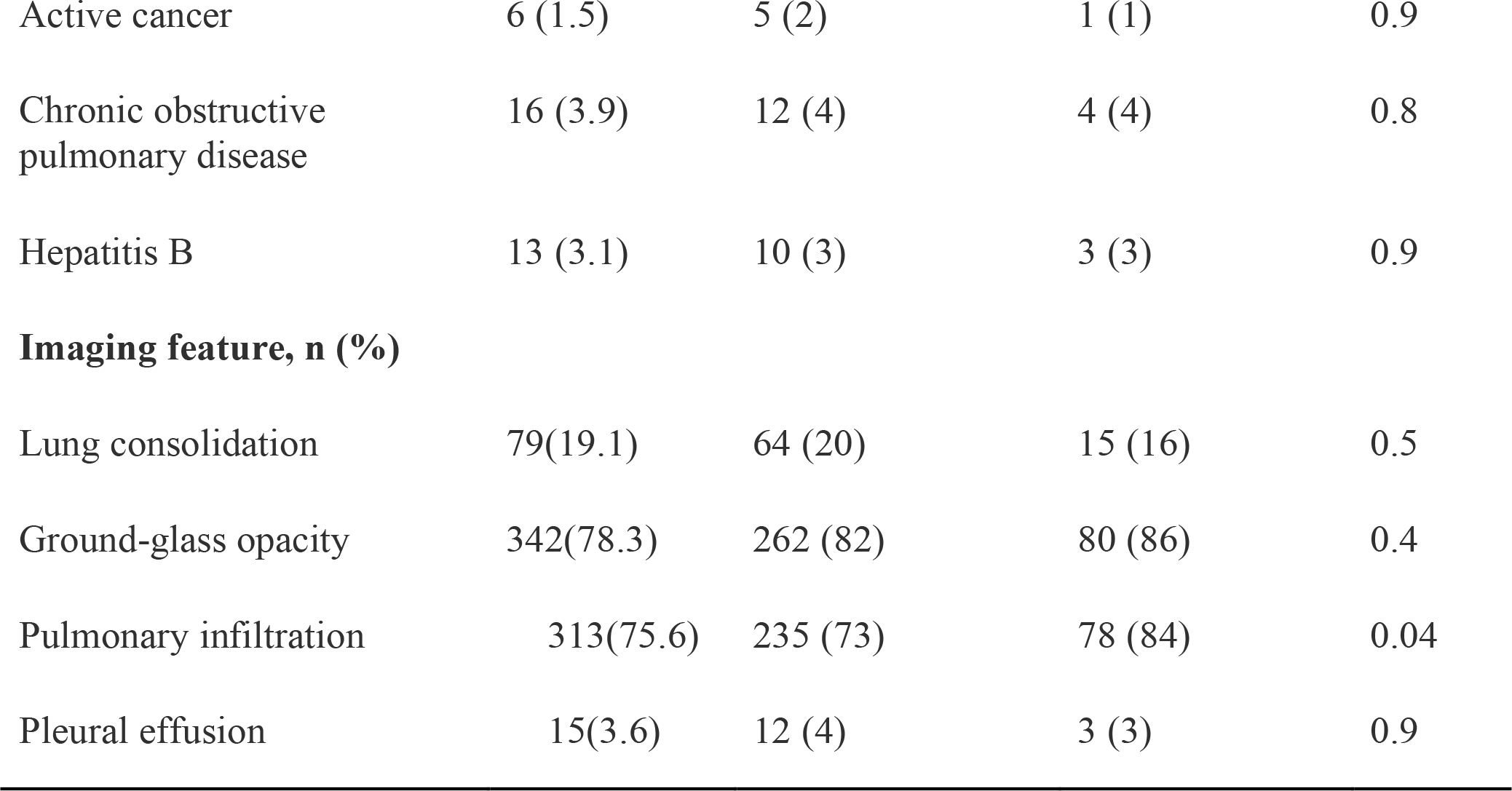
Demographics and baseline characteristics of COVID-19 patients with and without recurrence of SARS-CoV-2 PCR positivity. Data are n (%), n/N (%), mean (SD)

**Table 2:** Medication treatments of COVID-19 patients with and without recurrence of SARS-CoV-2 PCR positivity during the 1st hospitalization. The p-value is calculated based on the binomial test with medication treatment 6 (Lopinavir/Ritonavir/Arbidol) as the active control.

| **Medication Treatment\*, n (%)** | **No-recurrence, N=321** | **Recurrence, N=93** | **Sample Size, N=414** | **Recurrence Probability** | **P-value** |
| --- | --- | --- | --- | --- | --- |
| **6** | 25 (8) | 9 (10) | 34 (8) | 0.290 | NA |
| **5** | 25 (8) | 9 (10) | 34 (8) | 0.265 | 0.406 |
| **56** | 88 (27) | 28 (30) | 116 (28) | 0.241 | 0.099 |
| **156** | 39 (12) | 6 (6) | 45 (11) | 0.133 | 0.008 |
| **356** | 26 (8) | 5 (5) | 31 (7) | 0.161 | 0.063 |
| **456** | 28 (9) | 6 (6) | 34 (8) | 0.176 | 0.079 |
| **156+** | 30 (9) | 7 (8) | 37 (9) | 0.189 | 0.095 |
| **the rest** | 63 (20) | 23 (25) | 86 (21) | 0.267 | 0.298 |

\*1 = Methylprednisolone, 2 = Tocilizumab, 3 = Oseltamivir, 4 = Ribavirin, 5 = Interferon, 6 = Lopinavir/Ritonavir/Arbidol, 7 = Favipiravir, 8 = Chloroquine. Drug combo 156+ includes: 123456, 12356, 12456, 1256, 13456, 1356, 13568, 1456, 1567, 1568. Drug combo the rest includes: 0 (non-drug taken), 1, 135, 136, 146, 15, 157, 16, 345, 346, 35, 3568, 357, 36, 38, 4, 45, 4568, 46, 567, 568, 57, 58, 8.

**Figure 1:**
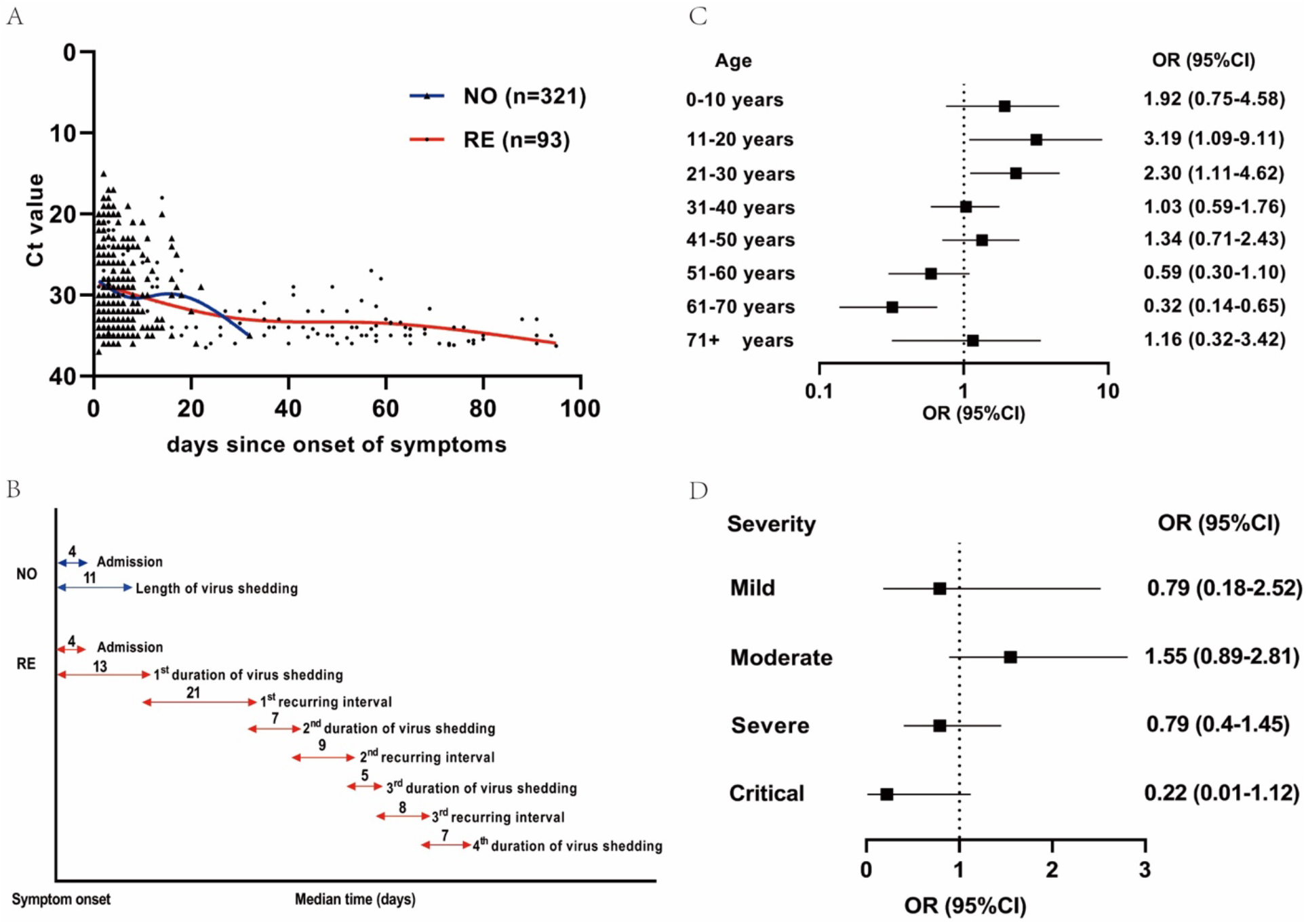
Temporal patterns of viral shedding, and association of recurrence with age and disease severity. Viral load [threshold cycle (Ct) values] detected by qRT–PCR in NP swabs from patients infected with SARS-CoV-2. A. The detection limit was Ct = 40, which was used to indicate negative samples. The thick lines show the trend in viral load during the whole course, using smoothing splines for NO and RE groups. B. The Covid-19 timeline summarizes the median duration (days) from the onset of symptoms to recurrence of NP swab PCR positivity. C. Association between age and recurrence. Age was depicted in deciles. D. Association between disease severity and recurrence. Disease severity is divided into four categories. qRT-PCR denotes real-time quantitative polymerase chain reaction; NP, nasopharyngeal; SARS-CoV-2, Severe acute respiratory syndrome coronavirus 2; NO, non-recurring patients; RE, recurring patients; Covid-19, Coronavirus disease 2019; OR (95% CI) referred to odds ratios with the corresponding 95% confidence intervals. The 95% CIs were not adjusted for multiple testing.

### Analysis of non-recurrence and recurrence

Table 1 and Supplemental Table 1 show the distribution of demographic characteristics and clinical information. There is no significant difference between recurring and non-recurring groups in terms of sex, disease conditions, comorbidity, imaging features, biochemical parameters and most of medical symptoms. Statistically significant differences between the two groups were observed for age and clinical symptoms (cough and sputum) at admission. For recurring patients, the majority (81%) were moderate (Supplemental Table 1). The whole duration of virus shedding could last for 3 months from symptom onset (Figure 1A). Initial virus loads of NP swab at admission were similar in the recurring and non-recurring patients (Supplementary Table 1). Data were analyzed according to age decile and disease severity (Figure 1 C, D). Recurrence was more commonly seen in younger patients (P value < 0.001).

### Analysis of medication treatments effects in terms of recurring rate

Our analysis suggested that no individual drug had a significant impact on the recurring probability (Supplementary Table 1). Different drug combinations were also studied in this paper. The corresponding medication regimen is summarized in Table 2. 63% of patients received antivirus treatments based on combination of Interferon (Inhale), Arbidol and Lopinavir/Ritonavir. 28% of patients received Lopinavir/Ritonavir, Arbidol combined with Interferon. 11% of patients received Lopinavir/Ritonavir/Arbidol/Interferon combined with Methylprednisolone. 7% and 8% of patients received Lopinavir/Ritonavir/Arbidol/Interferon combined with Oseltamivir and Ribavirin respectively. The Bonferroni corrected binomial tests at alpha level 0.1 revealed that the recurring rate of Methylprednisolone/Interferon/Lopinavir/Ritonavir/Arbidol was significantly lower than the one under Lopinavir/Ritonavir/Arbidol alone.

The left panel of Figure 2 illustrates the age-specific BMI distribution of recurring (red dots) and non-recurring (black dots) groups treated by different drug combinations. Supplementary Table S6 shows that certain clinical factors have some serious discrepancies between drug treatments (e.g., the average ages of patients treated with drug combo 5 and 156+ is 42 and 56, respectively). If it is assumed that these predictors are important for drug selection, then it is very important to try to correct this imbalance. Therefore, a VT analysis was performed, and the simultaneous MCB 95% confidence sets across the eight drug treatments were constructed. The upper bound of drug combo 156 is smaller than 0 while the lower bounds of all other drug combos is larger than 0, suggesting that across the whole patients of 20 to 70 years old, drug combo 156 is the best treatment with the recurring rate at least on average 6.7% (95% CI: 5.5% to 8.1%) lower than other drug treatments under comparison.

**Figure 2:**
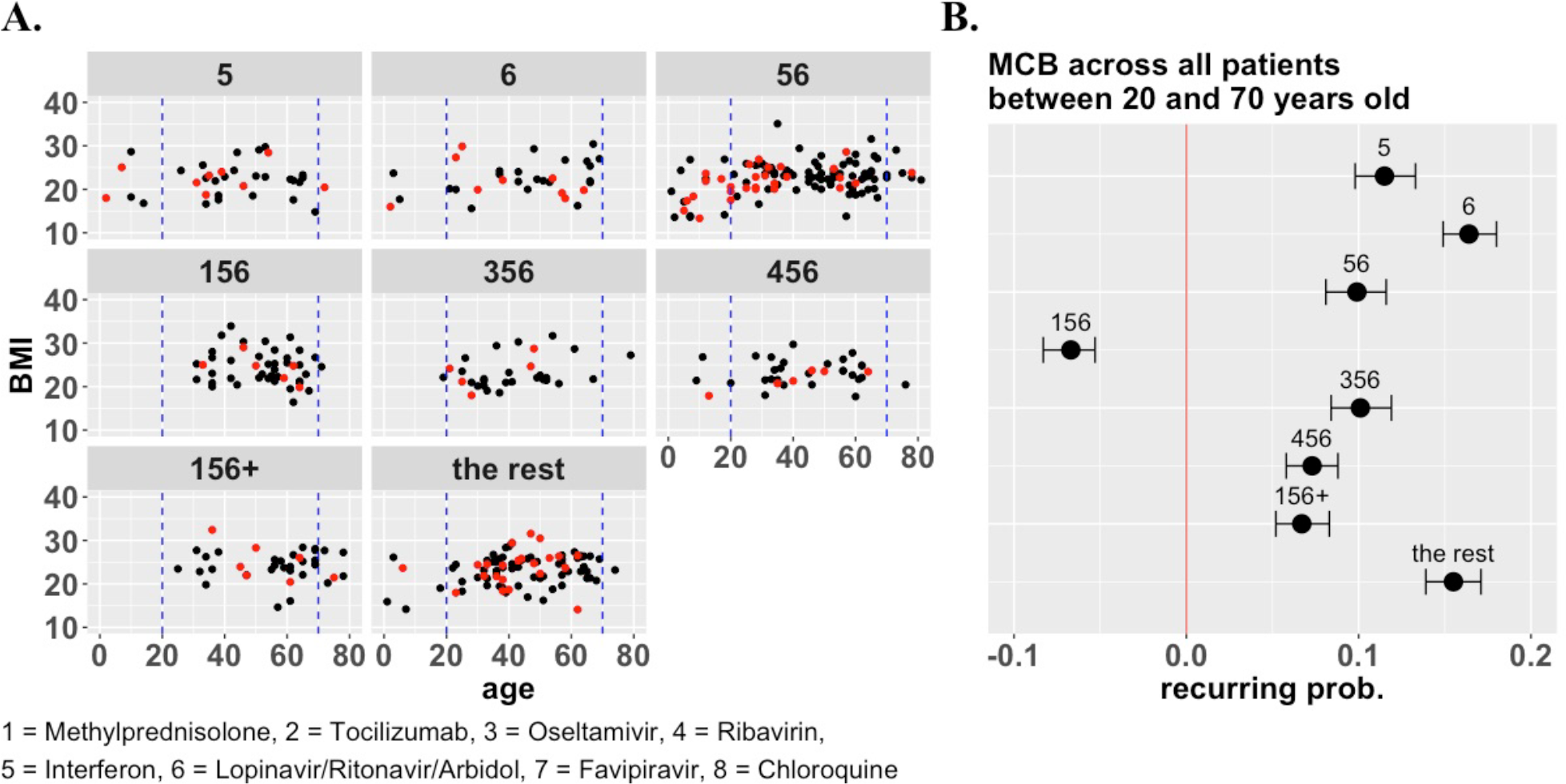
The effects of different drug treatments on SARS-CoV-2 recurrence. Eight medication treatment combinations (i.e., 5, 6, 56, 156, 356, 456, 156+, the rest) were administered to patients (Table 2). A. The age-specific BMI distribution of recurring (red dots) and non-recurring (black dots) groups treated by different drug combinations. B. MCB was performed on the recurring rate among all eight medication treatment groups, i.e., for each treatment group, its recurring rate was compared with the minimum recurring rate of all the other treatment groups. A simultaneous MCB 95% confidence set was constructed. The upper bound of drug treatment 156 is smaller than 0, which means drug treatment 156 is superior to other treatments.

Although the drug combo 156 is the best treatment overall (Figure 2), patients in different age groups and with different delay times of hospitalization may react differently under different drug treatments. The interaction among age, hospitalization delay and drug treatment on SARS-CoV-2 recurring rate is shown in Figure 3. A summary of drug treatments preference (in different subgroups stratified by age and hospitalization delay) is shown in Table 3. For example, our analysis predicted that the 156+ treatment administered to patients who are 20 to 27 years old with hospitalization delay greater than 5 days will result in the recurring rate at least on average 4.2% (95% CI: 0.4% to 6.3%) lower than the other 7 drug treatments. Therefore 156+ treatment is recommended. On the other hand, there were no significant differences in recurring rates among 156, 456, and 156+ for patients who are 27 to 31 years old with hospitalization delay greater than 5 days. Therefore all three treatments are superior to others.

**Figure 3:**
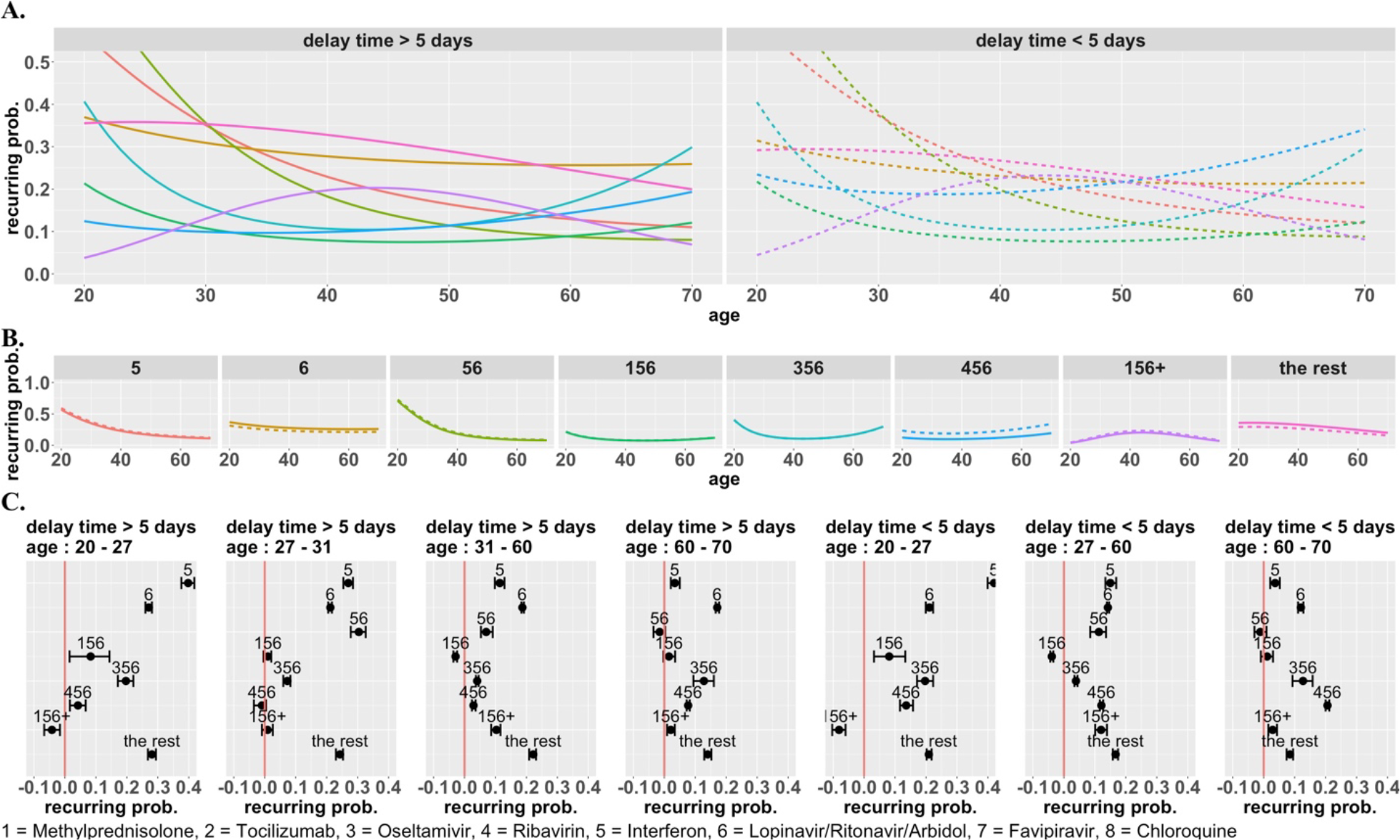
Age and hospitalization delay has important impacts on medical treatment of SARS-CoV-2 recurrence. A. The interaction among age, hospitalization delay and drug treatment on SARS-CoV-2 recurring rate. The predicted curves were generated by the Beta regression, and stratified by hospitalization delay. B. Same predicted curves stratified by eight medication treatment combinations, instead of hospitalization delay. C. MCB was performed within different patient subgroups stratified by age and hospitalization delay. If there is only one drug treatment whose upper bound is less than 0, then that treatment outperforms all the other treatments; on the other hand, if there are multiple drug treatments whose confidence intervals cover zero, then there is no significant difference between these drug treatments, and they are superior to others.

The interaction among age, obesity and drug treatments on recurring rate is shown in Figure 4. Overweight patients between 20 to 70 years old showed a totally different pattern from those with normal weights under different treatments: for overweight patients, drug combo 456 is preferred for patients who are 20 to 50 years old; drug combo 56 is preferred for patients who are 50 to 70 years old.

**Figure 4:**
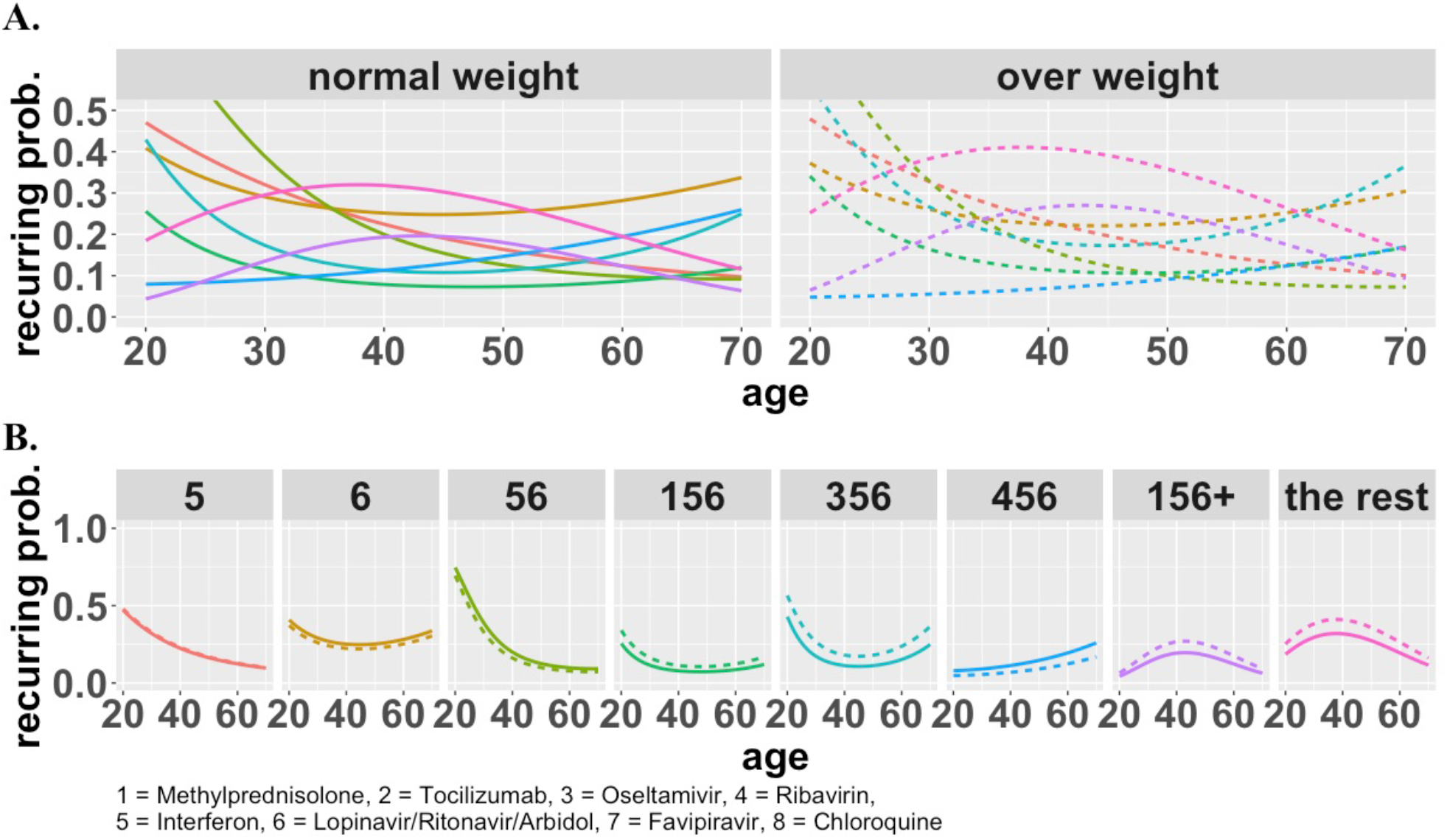
Obesity has important impacts on medical treatment of SARS-CoV-2 recurrence. A. The interaction among age, obesity and drug treatment on SARS-CoV-2 recurring rate. The predicted curves were generated by the Beta regression, and stratified by obesity. Patients between 20 to 70 years old with over weights showed a totally different pattern from those with normal weights under different treatments: for overweight patients, drug combo 456 is preferred for patients who are 20 to 50 years old; drug combo 56 is preferred for patients who are 50 to 70 years old. B. Same predicted curves stratified by eight medication treatment combinations, instead of obesity.

## Discussion

Our aforementioned statistical analyses on this one medical center real-world data revealed that different drug combinations could lead to different recurring rates, with the combination of Methylprednisolone/IFNα/Lopinavir/Ritonavir/Arbidol having the overall lowest recurring rate. In our study, we found that each age group has different optimal drug combinations. For the younger group (age 20-27), Methylprednisolone/IFNα/Lopinavir/Ritonavir/Arbidol with more drug combinations, such as Oseltamivir or Ribavirin, led to the best outcomes, while as for the older group (age 60-70), IFNα/Lopinavir/Ritonavir/Arbidol, had optimal outcomes. Interestingly, for the obese COVID-19 patients, the combination of Ribavirin/IFNα/Lopinavir/Ritonavir/Arbidol led to lowest recurring rate for age group of 20-50 year, while as in the age group of 50-70, the drug combination of Interferon/Lopinavir/Ritonavir/Arbidol resulted in a lower recurring rate. In the drug combination of Lopinavir/Ritonavir/Arbidol, only 20% patients took Arbidol in the group, and therefore, we think the effect mainly come from Lopinavir/Ritonavir. Our study can provide clues for better personalized medication treatments and potentially save significant medical resources in re-admission of COVID-19 patients.

In this paper, we applied SMOTE to address the issue of class imbalance (i.e., rare event), and performed VT analysis to minimize the effect of confounding clinical factors, while preserving the distribution of recurring and non-recurring groups under each drug treatment (Supplementary Table S4). To identify the best treatment that minimizes recurring probability, it is natural to consider post-hoc all pairwise comparisons. However, if two treatments are ineffective to the extent that neither of them can minimize the recurring probability, then it is not of primary interest to know which one of those two treatments is better and the inference that neither is the best suffices. We therefore proposed MCB to select the best drug treatment. Another essential advantage of MCB is that, since MCB is based on the simultaneous confidence set, instead of the p-value, it provides additional information of effective size and variation.

It is still inconclusive that the recurrent patients are transmissible or not, although from other virus experiences such patients are unlikely transmissible(9). There is still no firm evidence yet that immunity developed at the first infection is fully protective and antibody titers are associated with protection from reinfection. It has been suggested that COVID-19 recurrence may be a resurgence, not reactivation, depending on different tests and each person’s immunity(10). This idea is supported by the previous reported case that a patient ready-for-discharge after three consecutive negative nasopharyngeal swabs died of cardiac arrest. The SARS-COV-2 virus RNA was detected in the lung tissue by qPCR and SARS-CoV-2 nucleocapsid was detected by immunohistochemical (IHC) staining in the lung tissue. In addition, due to a large of a potential asymptomatic transmission of SARS-CoV-2, it is not clear that the COVID-19 recurrent patients have transmissible or not (8, 11).

These results may reflect the pathological characteristics of COVID-19. In our analysis, IFNα has been the main regiment in all good performance groups of drugs. This is not too surprising as IFNs is induced by most viruses and a major antiviral cytokine for multiple types of viruses(12-14). First, many viruses, such as Ebola, CMV, and EMCV, utilize SUMOylation to inhibit anti-viral intrinsic and innate immunity, such as reducing IFNα production and inhibiting STAT1/3, PML, IRFs, and NFκB(15-19). The Ebola viral VP35 interacts with IFNα regulatory factors, IRF3/7 and PIAS1, resulting in the SUMOylation and their transcriptional repression, therefore, suppresses the production of type I interferons(20). Therefore, it has been controversial in clinic whether IFNα alone should be used for treating high pathological viruses, such as SARS-CoV-2, SARS and MERS, although IFNαs were empirically recommended as one of the therapeutic options for COVID-19 in clinical practice(21). In both animal studies and clinic, the early treatment of IFN rescued mice from lethal doses of SARS-CoV and MERS, however, late IFN administration delayed viral clearance and exacerbate immunopathology(12). Another study has also shown IFN-I response timing relative to virus replication determined MERS coronavirus infection outcomes(13). Indeed, our studies suggest that treatment of moderate to severe COVID-19 patients with IFNα alone can ameliorate clinical outcomes. Significant clinical improvements and virus nucleotide transition to negative have been observed in our clinical trial for moderate to severe COVID19 patients using an engineered IFNα, rSIFN-co, strongly suggesting that IFNα is antagonized by viruses is critical for host immune response against viral infection(Li, C et al., submitted). This result may confirm the nature of SARS-CoV-2 and related virus infections, such as SARS and MERS, to dysregulation of IFNα induction at early stage of infection(14).

Severe COVID-19 patients often develop acute respiratory distress syndrome (ARDS) or secondary haemophagocytic lymphohistiosus(sHLH)(15, 16). Both ARDS and sHLH are hallmarks of overwhelmed cytokine productions, so called cytokine storm or cytokine release syndrome (CRS), which is one of main causes of mortality(17),(18). In supporting the notion of anti-cytokine storm may be beneficial to COVID-19 patients, the administration of anti-inflammation drug, Methylprednisolone, slowed down the disease progress and reduced death rate(19).Findings from a preliminary, uncontrolled study revealed that IFN alfacon-1 plus corticosteroids was associated with reduced disease-associated impaired oxygen saturation, more rapid resolution of radiographic lung abnormalities in SARS patients, demonstrating antiviral activity against MERS(18, 19).

It was initially reported that the HIV protease inhibitors, Lopinavir–Ritonavir, had no benefit for COVID-19 patients as compared with standard care in terms of clinical improvement and mortality(22). But several responses pointed out that the conclusion was made prematurely because of small sample size and too late for treatment during COVID-19 disease progression(23). Lopinavir, was discovered to be active against SARS-CoV, the virus causing SARS in 2003 in an in vitro screening, and later, was shown together with Ritonavir to reduce ARDS and mortality rate in humans with SARS^19^. Case reports also suggested that the triple combination of Lopinavir–Ritonavir with IFNα or β improved virus clearance and survival(24-26). Our discovery also supports a recently a clinical trial program, called SOLIDARITY, launched by the WHO, aiming to quickly repurpose known or clinical drugs for CIVID-19 pandemic, has one arm of triple combination, Lopinavir–Ritonavir with IFNβ(27). However, our former Covid-19 study demonstrated that lopinavir-ritonavir-induced liver injury was characterized as a high level of total bilirubin and gamma-glutamyl transferase, and was an independent risk factor (odds ratio, 4.4; 95% confidence interval, 1.5-13.17) of liver injury during hospitalization(28).

The SARS-CoV-2 mimics the influenza virus in clinical presentation, transmission mechanism, and seasonal coincidence. A recent study reports four co-transfection cases with the SARS-CoV-2 and influenza virus, highlighting the urgent need for precision diagnosis and treatment for the co-infection(26, 27). In the last pandemic of the SARS in 2003, patients with fever, cough or sore throat had a 5% of influenza virus positive rate(29). This raises the concerns that there might be mixed infections of seasonal influenza and the SARS-CoV-2. Interestingly, we found out that the combination of anti-influenza virus drug, oseltamivir, with Interferon/Lopinavir/Ritonavir/Arbidol, has very good outcome (recurring rate of 0.172), supporting the hypothesis of co-infection of influenzas and SARS-CoV-2. Especially the current COVID-19 pandemic is overlapping with the flu season. This suggests that we need to screen the people with fever, cough or sore throat for both viruses with oral, NP and even anal swabs in respiratory infectious diseases surveillance systems.

### Limitation

In this cohort study during the COVID-19 emergency situation, the drugs and drug combinations are more than usual medical practice. Many medications were applied by previous literatures or recommendations of Chinese Health Committee guidelines. The safety concerns of using multiple drugs (>3) need more careful evaluations in the future. The study is a retrospective study with no control group in each treatment. The sample sizes in each group vary and are not equally distributed, which is a challenge for analysis. In addition, the recurring patients are still transmissible or not is still inconclusive. Some effective drugs, such as Remdesivir, were not included in this study.

## Method

### Study design and participants

The cohort included consecutive PCR confirmed Covid-19 patients who were admitted to The Second Affiliated Hospital of Southern University of Science and Technology, Shenzhen, China between January 11, 2020 to February 16, 2020 and completed their hospital course (discharge alive) by March 26, 2020. The latest follow-up date was May 5, 2020. All discharged COVID-19 patients were subject to strict quarantine at home or a designated center for 4 weeks. Regular follow-up was performed every 3-5 days during the quarantine.

### Data collection

Methods for laboratory confirmation of SARS-CoV-2 infection have been described in our previous study(28). Diagnosis, disease severity and treatment for COVID-19 infection were based on the preliminary diagnosis and treatment protocols (6th edition) from the National Health Commission of China (National Health Commission of the People’s Republic of China and National Administration of Traditional Chinese Medicine. Diagnosis and Treatment Scheme for Novel Coronavirus Pneumonia (Trial). 2020 [Feburary 19, 2020]. The sixth edition.:Available from: http://www.nhc.gov.cn/xcs/zhengcwj/202002/8334a8326dd94d329df351d7da8aefc2.shtml. The following information was collected from each patient: epidemiologic and demographic data, underlying diseases, clinical characteristics and laboratory testing at admission, medication regime during the hospitalization. Clinical characteristics, laboratory findings and images were also collected during the follow-up. Medication included Methylprednisolone, Tocilizumab, Oseltamivir, Ribavirin, Interferon (inhale), Lopinavir/ritonavir, Arbidol, Favipiravir and Hydroxychloroquine. Routine blood examinations were complete blood counts, serum biochemical tests (including renal and liver function, creatine kinase, infection-related biomarkers, coagulation function). Chest x-ray or CT scans were done for all patients. Early morning NP swabs were analyzed every 3-5 days till quarantine ends.

### Definitions

Disease severity were categorized into mild, moderate, severe and critical levels according to the protocols from the National Health Commission of China. Discharge criteria included improvement of respiratory symptoms and radiological lesions, two consecutive negative NP swab tests sampled >1 day apart. Recurrence of positive SARS-CoV-2 RNA test referred to a positive NP swab test emerging >3 days from the latest negative test in recovering patients. Recurring interval referred to the time slot from the last negative NP test to positivity recurrence. The primary endpoint was recurrence of positive NP swab testing of patients within 28-day quarantine. Patients who had recurrence of positive NP swab testing were defined as recurrence. Patients who did not have positive results within quarantine were treated as non-recurrence. We analyzed the profile, medication regimen and outcomes of patients between the recurring group and non-recurring group.

### Statistical Analysis

Among the 414 hospitalized patients who met the inclusion criteria for this study, there were a limited number of patients who were younger than 20 years old (34 patients), or older than 70 years old (16 patients). Therefore, our analysis was restricted to the age from 20 to 70 years old.

For predicting recurring probability of a given patient, we controlled for his/her confounding clinical factors, including age, BMI, gender, disease severity, hospitalization delay, consolidation, GGO, pulmonary infiltration, and pleural effusion. Since no individual drug had a significant impact on the recurring probability, eight medication treatment combinations were administered to patients in this study. We took the treatment of *Lopinavir/Ritonavir/Arbidol* as an active control because its effect for COVID-19 treatment was not so effective just by themselves(22). The Bonferroni corrected binomial test was performed for groups under the other drug treatments to test if the corresponding recurring probability was significantly lower than the one under the active control.

To minimize the effect of confounding clinical factors, a Virtual Twins (VT) analysis was performed in this study(30). Specifically, a separate random forest was fitted to each of the eight treatment combinations, and used to predict recurring probability for each patient in the corresponding treatment group. The forest was built on all confounding clinical factors: age, BMI, gender, disease severity, hospitalization delay, consolidation, GGO, pulmonary infiltration, and pleural effusion. Age, BMI, and hospitalization delay were ranked on average the top three most important predictors by all forests.

It is worth noting that within certain age groups, the recurring rate could be very small, and thus the recurring event can be considered as a rare event in those age groups. Random forest with classification trees might locally underestimate the recurring rate. Therefore, before conducting VT analysis, we applied the Synthetic Minority Oversampling Technique (SMOTE) to synthetically create additional observations by oversampling the recurring events through bootstrapping and k-nearest neighbors(31).

We then performed Multiple Comparisons with the Best (MCB) on the recurring rate among all eight treatment groups, i.e., for each treatment group, its recurring rate was compared with the minimum recurring rate of all the other treatment groups(32). A simultaneous 95% MCB confidence set was constructed. If the upper bound of a drug treatment’s recurring rate is smaller than zero, then its recurring rate is significantly smaller than that of any other drug treatment. Conversely, if the lower bound of a drug treatment’s recurring rate is larger than zero, then its recurring rate is significantly larger than some other drug treatment(s). Otherwise, a drug treatment with lower bound smaller than zero and upper bound larger than zero suggests that its recurring rate is not significantly different from the minimal recurring rate of all other drug treatments.

To evaluate the effects of age, BMI and hospitalization delay on the recurring probability under each drug treatment, we performed Beta regression analysis(33). Stratified by these three clinical factors, the best drug combination within each subgroup was determined by the MCB. A simultaneous 95% MCB confidence set was also constructed. To assess the performance of the prediction model in terms of the Area Under the ROC Curve (AUC), we performed Monte-Carlo cross-validation. That is, we created 1000 random splits of the dataset into training (4/5) and testing (1/5). For each such split, the model is fit to the training data, and the predictive accuracy is assessed using the testing data(34).

All statistical analyses were done with R version 3.6.3. The workflow of the whole statistical analyses procedure was available in Supplementary materials.

## Author contributions

JH conducted all the patient medical data analysis; SZ performed all statistical analysis;, FY, SW, MZ, TZ, LQ, ZX, and MY collected and organized the patient data; GW and VM read and edited the manuscript;, LL supervised the medical treatment and data collection; XC supervised the statistical analysis and manuscript writing and revising; JL designed, supervised the study, and manuscript writing.

## Data Availability

All the data are available upon request, except patient identity information.

## Acknowledgments

We would like to thank University of California, Riverside. Grant Number: AES-CE RSAP A01869 for Cui; UCR Academic Award to Jiayu Liao; and National Natural Science Foundation of China (No. 81501651 to Jia Huang

## Supplement Figures

**Figure S1:**
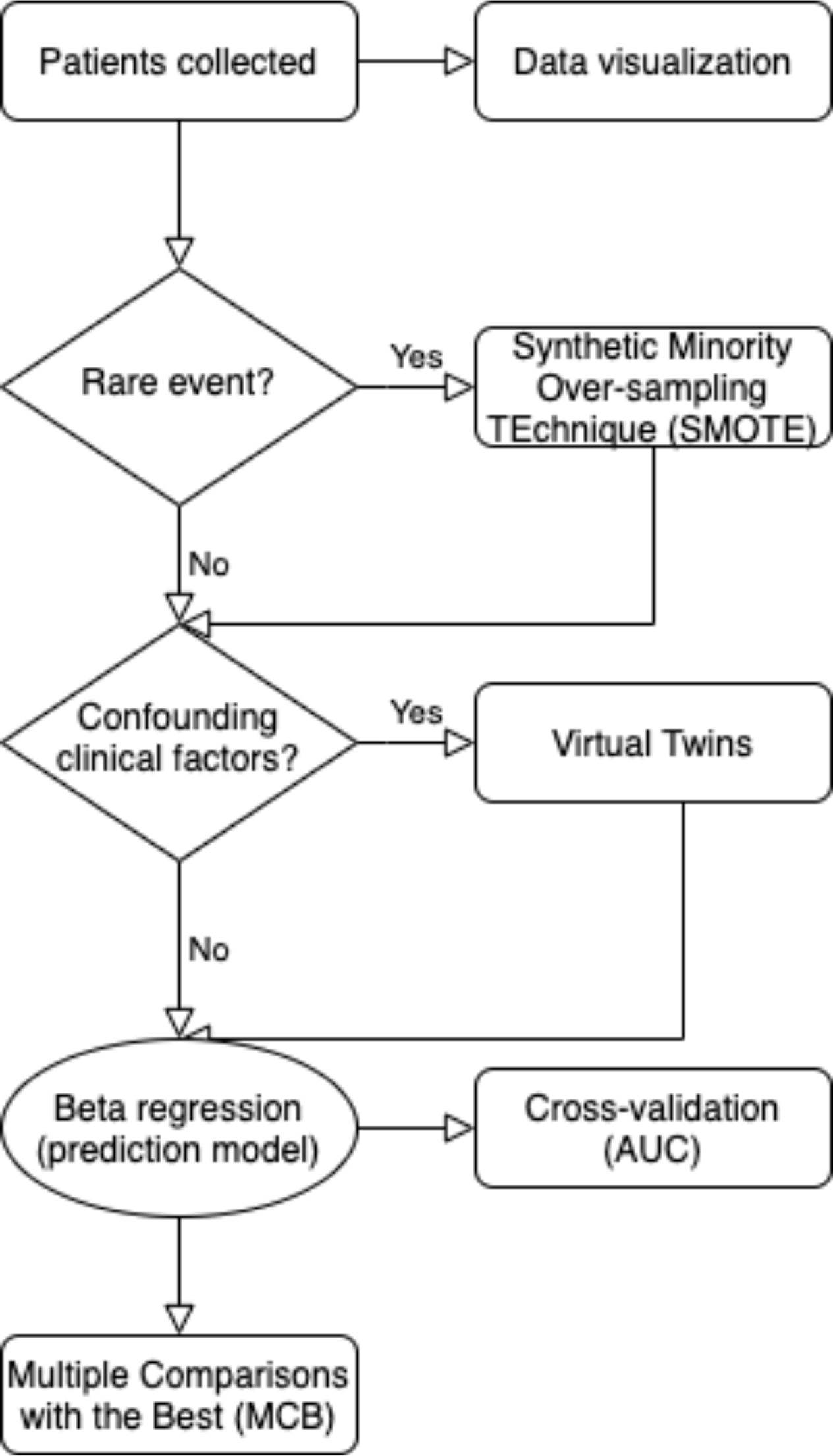
Workflow of statistical analysis.

**Figure S2:**
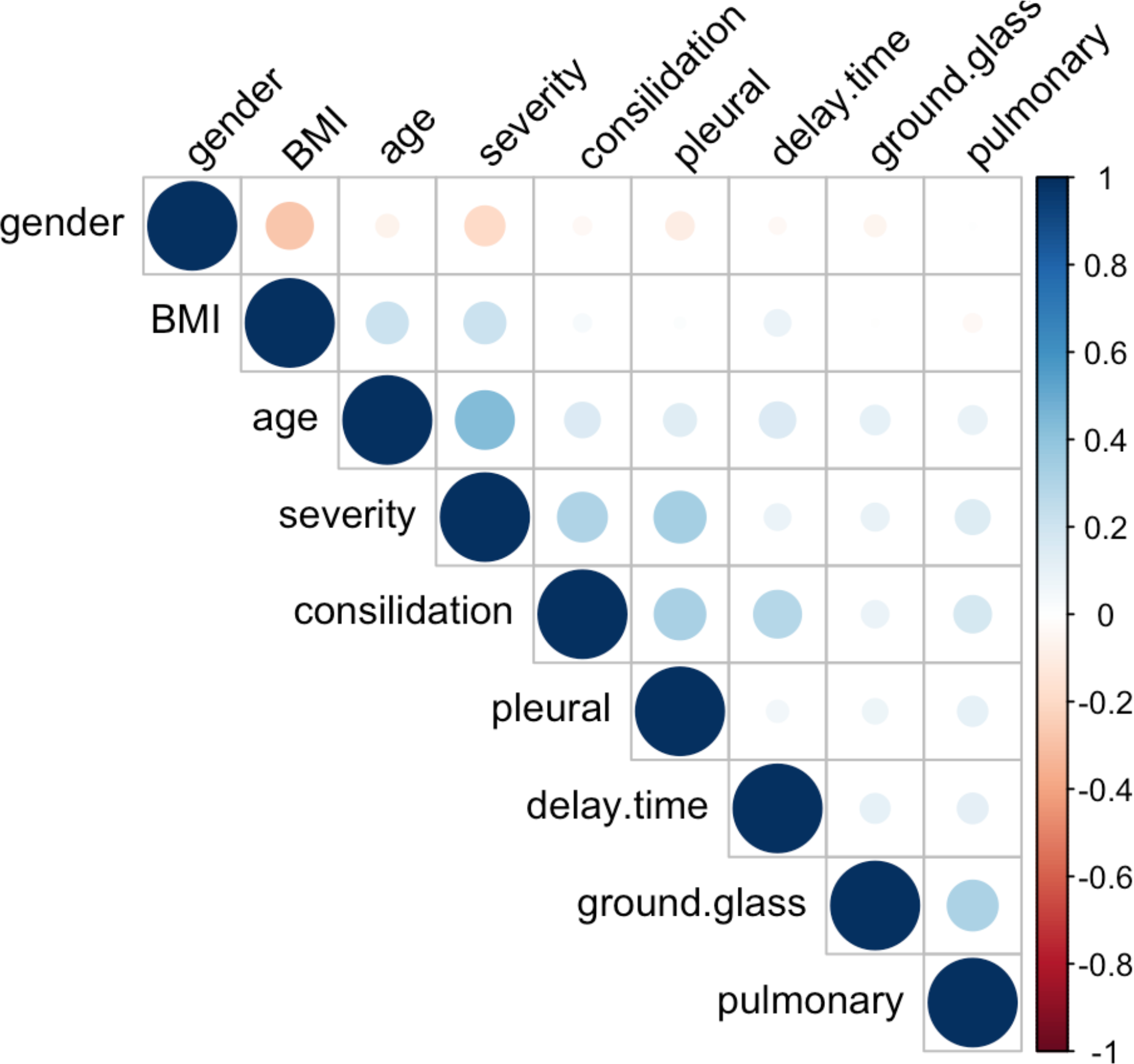
Pairwise correlations between clinical factors.

**Figure S3:**
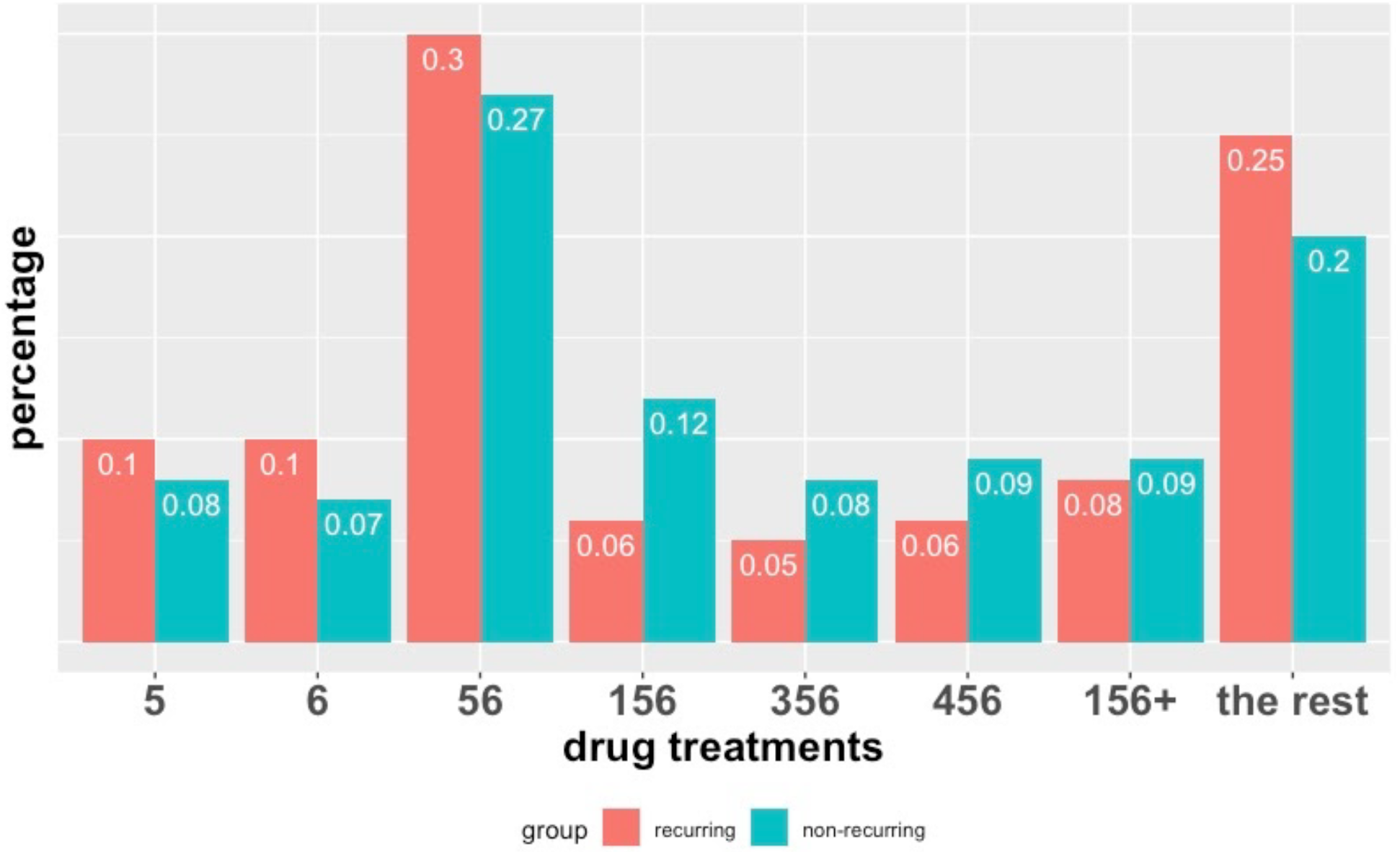
Drug treatment distributions (i.e., multinomial distribution) between recurring and non-recurring groups.

**Figure S4:**
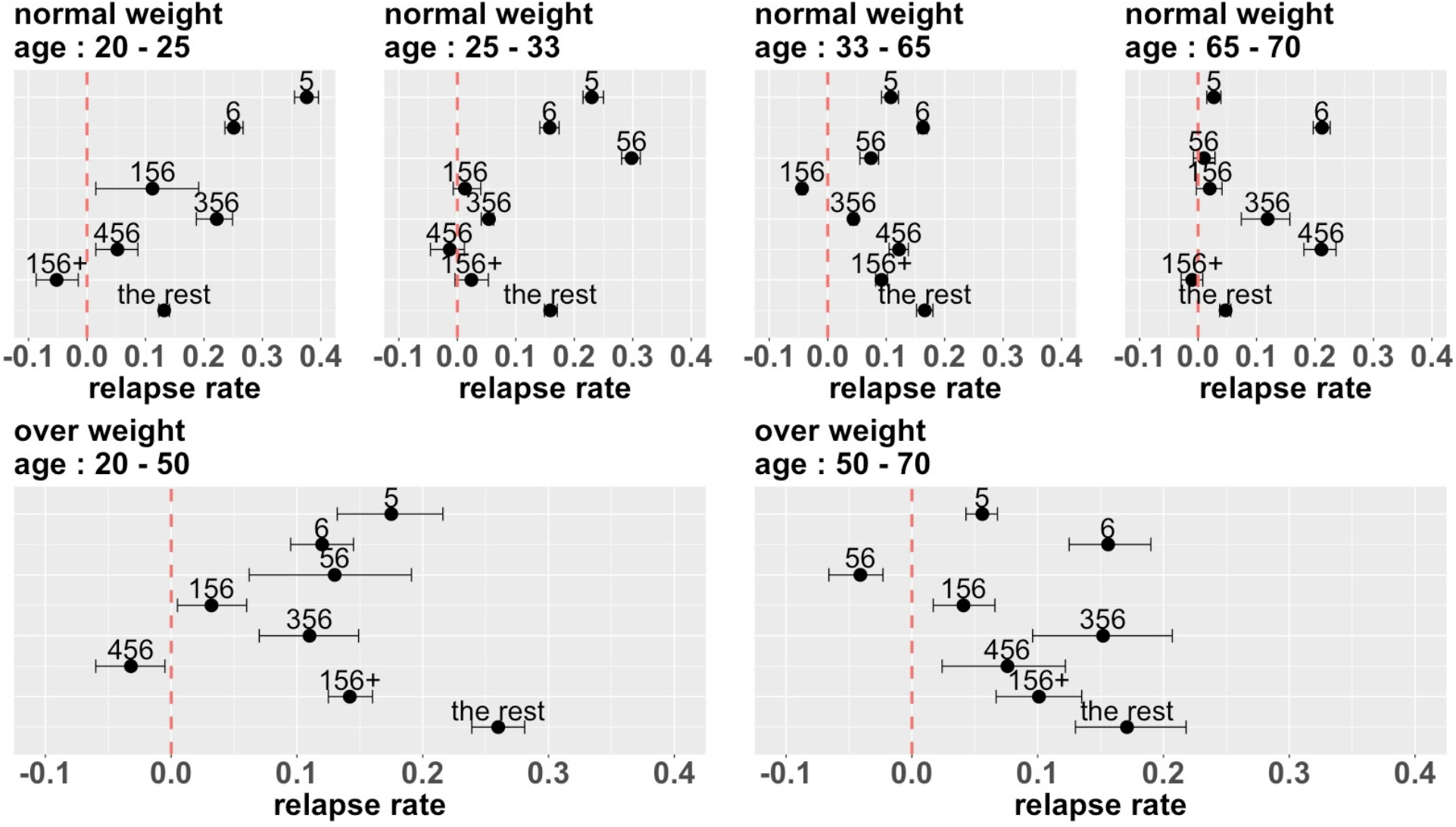
The impact of BMI on SARS-Cov-2 recurrence. The simultaneous MCB 95% confidence set was constructed within each subgroup stratified by age and obesity.

**Figure S5:**
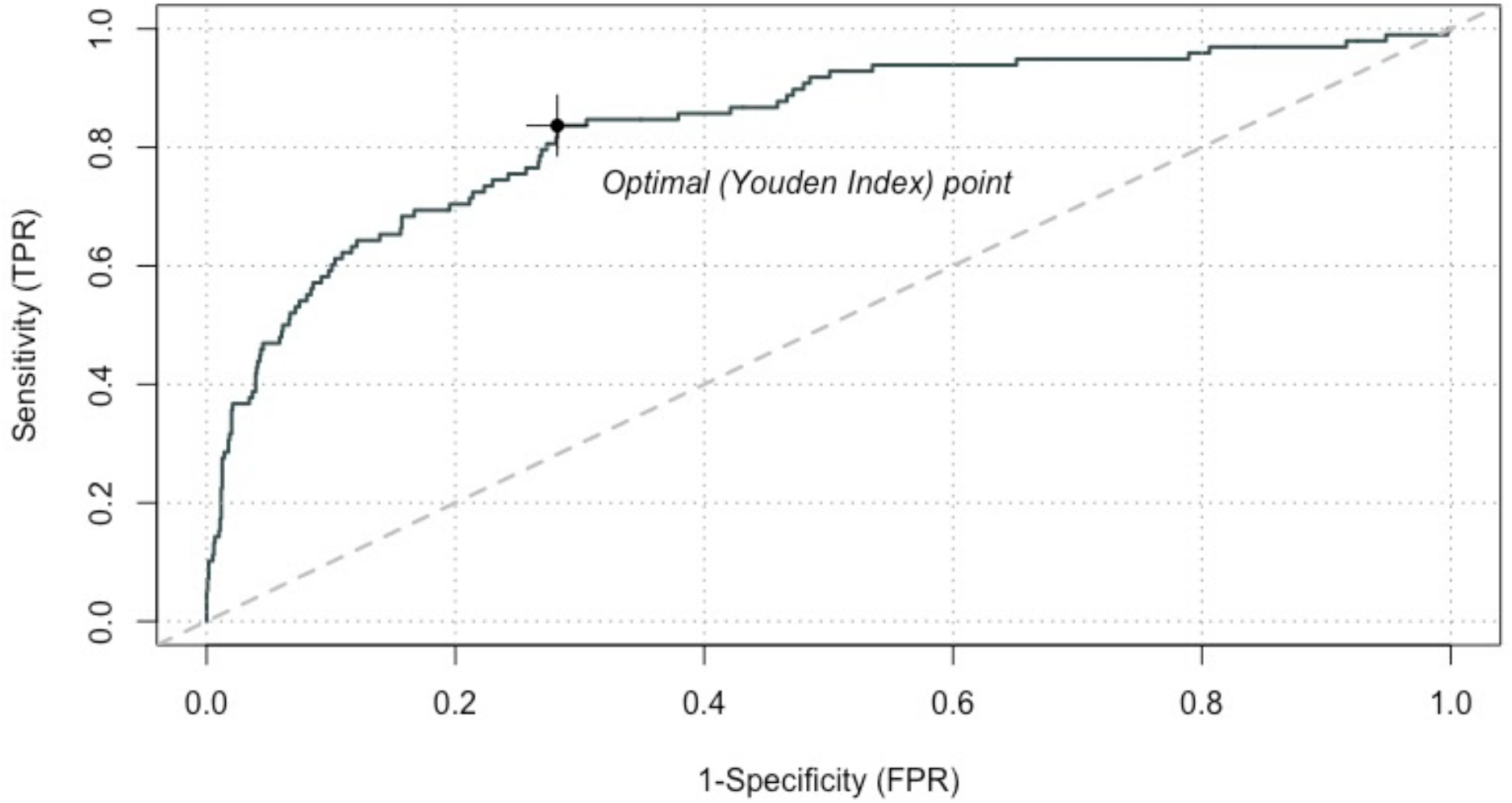
Assess the prediction model by cross-validation in terms of AUC. The average performance of the prediction model (i.e., AUC = 0.83). Statistics of AUCs: min = 0.75, median = 0.84, mean = 0.83, max = 0.89.

## Supplement Table

**Table S1:**
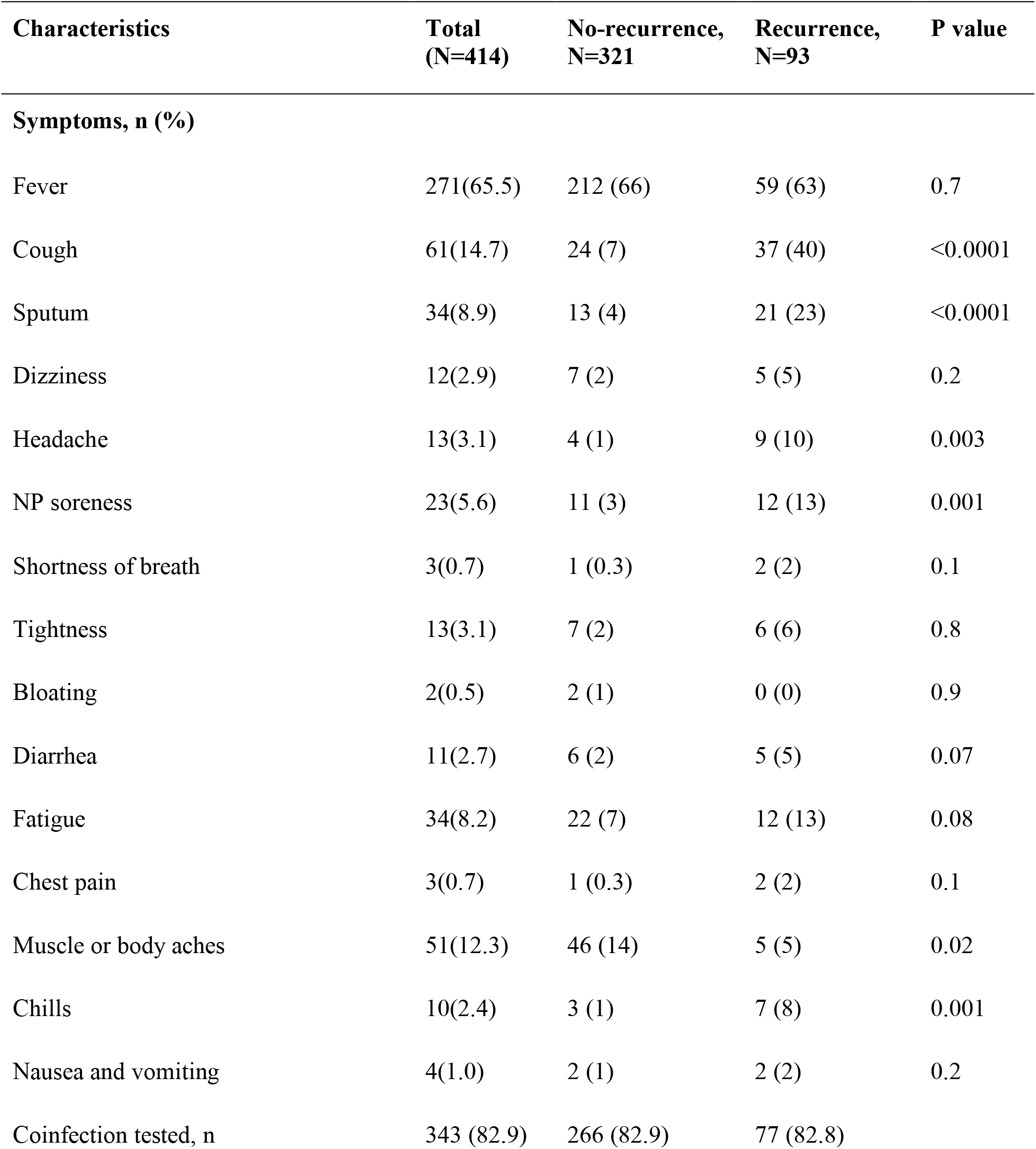
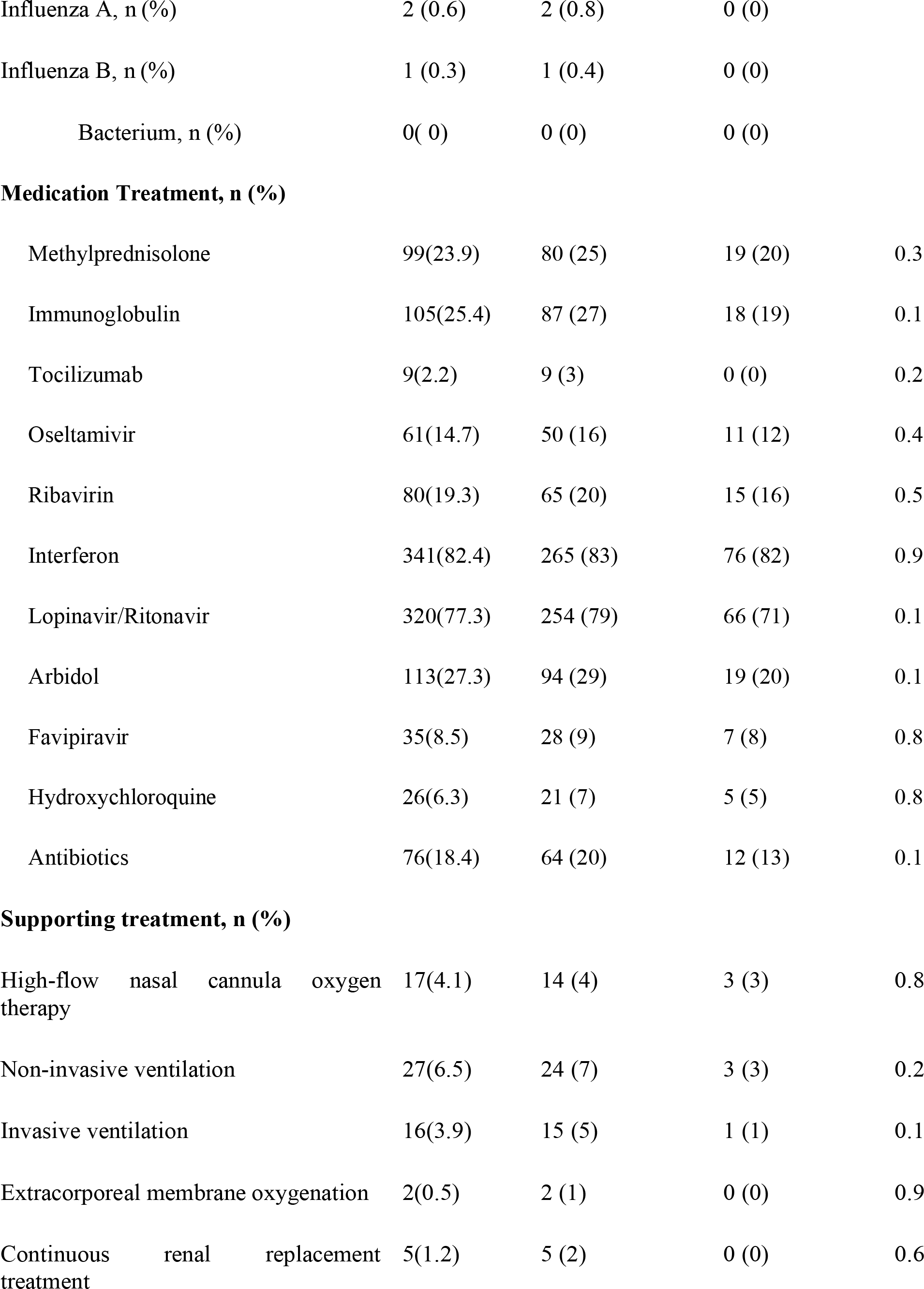
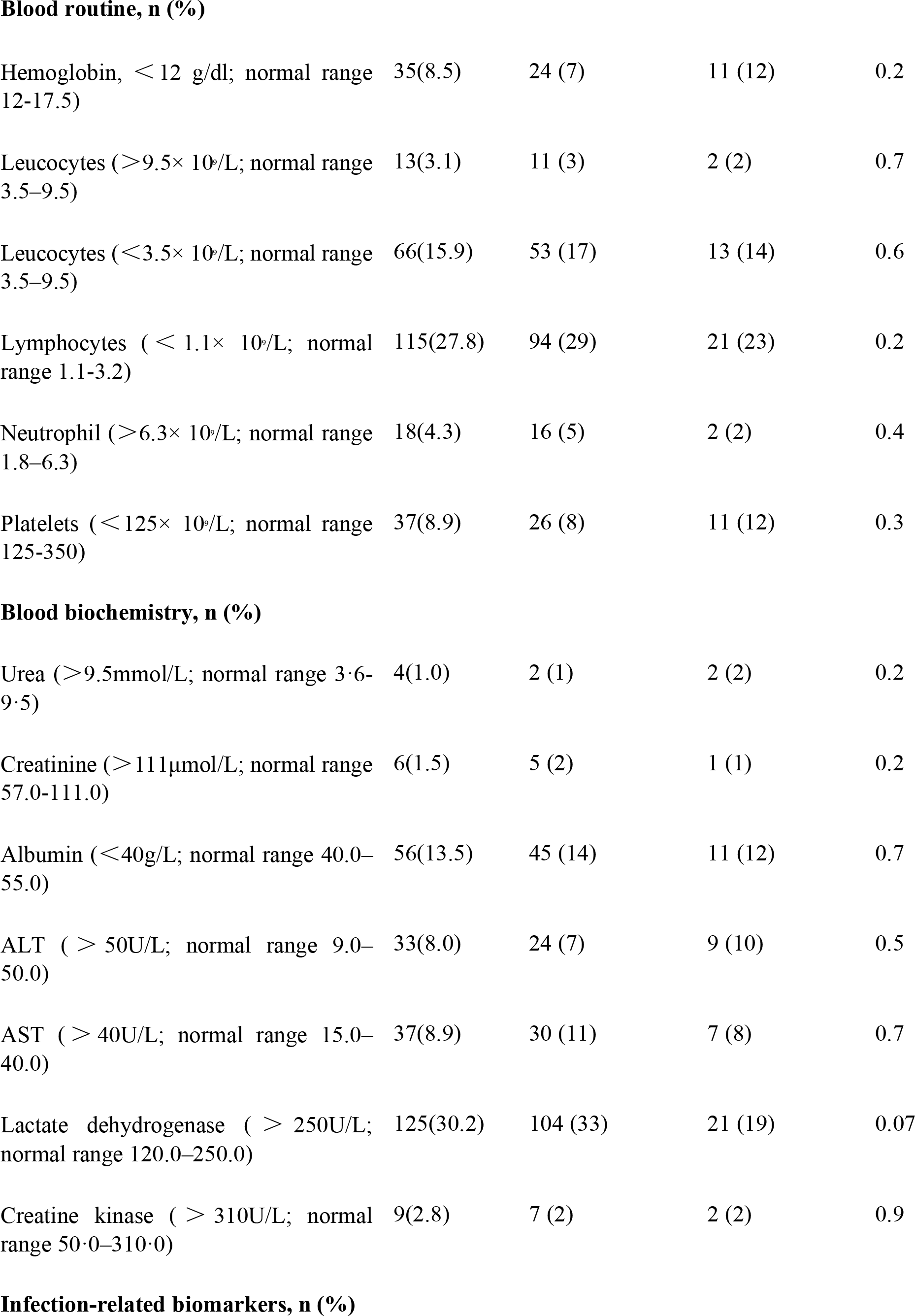
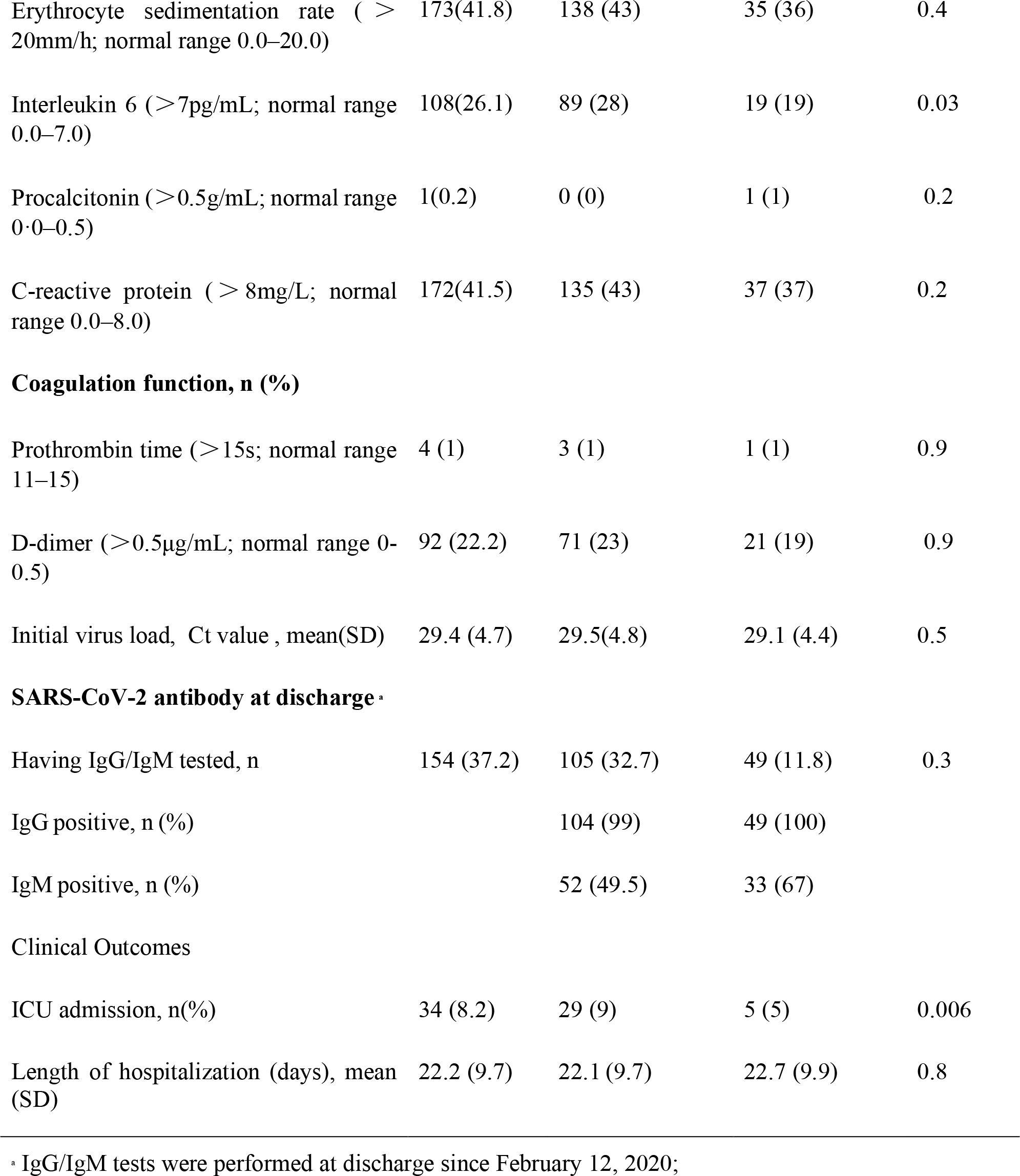
Clinical characteristics, laboratory findings, treatments, and outcomes of Covid-19 patients with and without recurrence of SARS-CoV-2 PCR positivity during hospitalization. Data are n (%), n/N (%), mean (SD). For statistical analyses, the Mann-Whitney U test was used to compare continuous variables and Fisher’s exact test was performed to compare categorical variables between groups.

| Characteristics | Total<br>(N=414) | No-recurrence,<br>N=321 | Recurrence,<br>N=93 | P value |
| --- | --- | --- | --- | --- |
| **Symptoms, n (%)** |  |  |  |  |
| Fever | 271(65.5) | 212 (66) | 59 (63) | 0.7 |
| Cough | 61(14.7) | 24 (7) | 37 (40) | <0.0001 |
| Sputum | 34(8.9) | 13 (4) | 21 (23) | <0.0001 |
| Dizziness | 12(2.9) | 7 (2) | 5 (5) | 0.2 |
| Headache | 13(3.1) | 4 (1) | 9 (10) | 0.003 |
| NP soreness | 23(5.6) | 11 (3) | 12 (13) | 0.001 |
| Shortness of breath | 3(0.7) | 1 (0.3) | 2 (2) | 0.1 |
| Tightness | 13(3.1) | 7 (2) | 6 (6) | 0.8 |
| Bloating | 2(0.5) | 2 (1) | 0 (0) | 0.9 |
| Diarrhea | 11(2.7) | 6 (2) | 5 (5) | 0.07 |
| Fatigue | 34(8.2) | 22 (7) | 12 (13) | 0.08 |
| Chest pain | 3(0.7) | 1 (0.3) | 2 (2) | 0.1 |
| Muscle or body aches | 51(12.3) | 46 (14) | 5 (5) | 0.02 |
| Chills | 10(2.4) | 3 (1) | 7 (8) | 0.001 |
| Nausea and vomiting | 4(1.0) | 2 (1) | 2 (2) | 0.2 |
| Coinfection tested, n | 343 (82.9) | 266 (82.9) | 77 (82.8) |  |
| Influenza A, n (%) | 2 (0.6) | 2 (0.8) | 0 (0) |  |
| Influenza B, n (%) | 1 (0.3) | 1 (0.4) | 0 (0) |  |
| Bacterium, n (%) | 0( 0) | 0 (0) | 0 (0) |  |
| **Medication Treatment, n (%)** |  |  |  |  |
| Methylprednisolone | 99(23.9) | 80 (25) | 19 (20) | 0.3 |
| Immunoglobulin | 105(25.4) | 87 (27) | 18 (19) | 0.1 |
| Tocilizumab | 9(2.2) | 9 (3) | 0 (0) | 0.2 |
| Oseltamivir | 61(14.7) | 50 (16) | 11 (12) | 0.4 |
| Ribavirin | 80(19.3) | 65 (20) | 15 (16) | 0.5 |
| Interferon | 341(82.4) | 265 (83) | 76 (82) | 0.9 |
| Lopinavir/Ritonavir | 320(77.3) | 254 (79) | 66 (71) | 0.1 |
| Arbidol | 113(27.3) | 94 (29) | 19 (20) | 0.1 |
| Favipiravir | 35(8.5) | 28 (9) | 7 (8) | 0.8 |
| Hydroxychloroquine | 26(6.3) | 21 (7) | 5 (5) | 0.8 |
| Antibiotics | 76(18.4) | 64 (20) | 12 (13) | 0.1 |
| **Supporting treatment, n (%)** |  |  |  |  |
| High-flow nasal cannula oxygen therapy | 17(4.1) | 14 (4) | 3 (3) | 0.8 |
| Non-invasive ventilation | 27(6.5) | 24 (7) | 3 (3) | 0.2 |
| Invasive ventilation | 16(3.9) | 15 (5) | 1 (1) | 0.1 |
| Extracorporeal membrane oxygenation | 2(0.5) | 2 (1) | 0 (0) | 0.9 |
| Continuous renal replacement treatment | 5(1.2) | 5 (2) | 0 (0) | 0.6 |
| **Blood routine, n (%)** |  |  |  |  |
| Hemoglobin, < 12 g/dl; normal range 12-17.5) | 35(8.5) | 24 (7) | 11 (12) | 0.2 |
| Leucocytes (>9.5× 10<sup>9</sup>/L; normal range 3.5–9.5) | 13(3.1) | 11 (3) | 2 (2) | 0.7 |
| Leucocytes (<3.5× 10<sup>9</sup>/L; normal range 3.5–9.5) | 66(15.9) | 53 (17) | 13 (14) | 0.6 |
| Lymphocytes ( < 1.1× 10<sup>9</sup>/L; normal range 1.1-3.2) | 115(27.8) | 94 (29) | 21 (23) | 0.2 |
| Neutrophil (>6.3× 10<sup>9</sup>/L; normal range 1.8–6.3) | 18(4.3) | 16 (5) | 2 (2) | 0.4 |
| Platelets (<125× 10<sup>9</sup>/L; normal range 125-350) | 37(8.9) | 26 (8) | 11 (12) | 0.3 |
| **Blood biochemistry, n (%)** |  |  |  |  |
| Urea (>9.5mmol/L; normal range 3·6-9·5) | 4(1.0) | 2 (1) | 2 (2) | 0.2 |
| Creatinine (>111μmol/L; normal range 57.0-111.0) | 6(1.5) | 5 (2) | 1 (1) | 0.2 |
| Albumin (<40g/L; normal range 40.0–55.0) | 56(13.5) | 45 (14) | 11 (12) | 0.7 |
| ALT ( > 50U/L; normal range 9.0–50.0) | 33(8.0) | 24 (7) | 9 (10) | 0.5 |
| AST ( > 40U/L; normal range 15.0–40.0) | 37(8.9) | 30 (11) | 7 (8) | 0.7 |
| Lactate dehydrogenase ( > 250U/L; normal range 120.0–250.0) | 125(30.2) | 104 (33) | 21 (19) | 0.07 |
| Creatine kinase ( > 310U/L; normal range 50·0–310·0) | 9(2.8) | 7 (2) | 2 (2) | 0.9 |
| **Infection-related biomarkers, n (%)** |  |  |  |  |
| Erythrocyte sedimentation rate ( > 20mm/h; normal range 0.0–20.0) | 173(41.8) | 138 (43) | 35 (36) | 0.4 |
| Interleukin 6 (>7pg/mL; normal range 0.0–7.0) | 108(26.1) | 89 (28) | 19 (19) | 0.03 |
| Procalcitonin (>0.5g/mL; normal range 0.0–0.5) | 1(0.2) | 0 (0) | 1 (1) | 0.2 |
| C-reactive protein ( > 8mg/L; normal range 0.0–8.0) | 172(41.5) | 135 (43) | 37 (37) | 0.2 |
| **Coagulation function, n (%)** |  |  |  |  |
| Prothrombin time (>15s; normal range 11–15) | 4 (1) | 3 (1) | 1 (1) | 0.9 |
| D-dimer (>0.5µg/mL; normal range 0-0.5) | 92 (22.2) | 71 (23) | 21 (19) | 0.9 |
| Initial virus load, Ct value , mean(SD) | 29.4 (4.7) | 29.5(4.8) | 29.1 (4.4) | 0.5 |
| **SARS-CoV-2 antibody at discharge<sup>a</sup>** |  |  |  |  |
| Having IgG/IgM tested, n | 154 (37.2) | 105 (32.7) | 49 (11.8) | 0.3 |
| IgG positive, n (%) |  | 104 (99) | 49 (100) |  |
| IgM positive, n (%) |  | 52 (49.5) | 33 (67) |  |
| **Clinical Outcomes** |  |  |  |  |
| ICU admission, n(%) | 34 (8.2) | 29 (9) | 5 (5) | 0.006 |
| Length of hospitalization (days), mean (SD) | 22.2 (9.7) | 22.1 (9.7) | 22.7 (9.9) | 0.8 |

<sup>a</sup> IgG/IgM tests were performed at discharge since February 12, 2020;

**Table S2:**
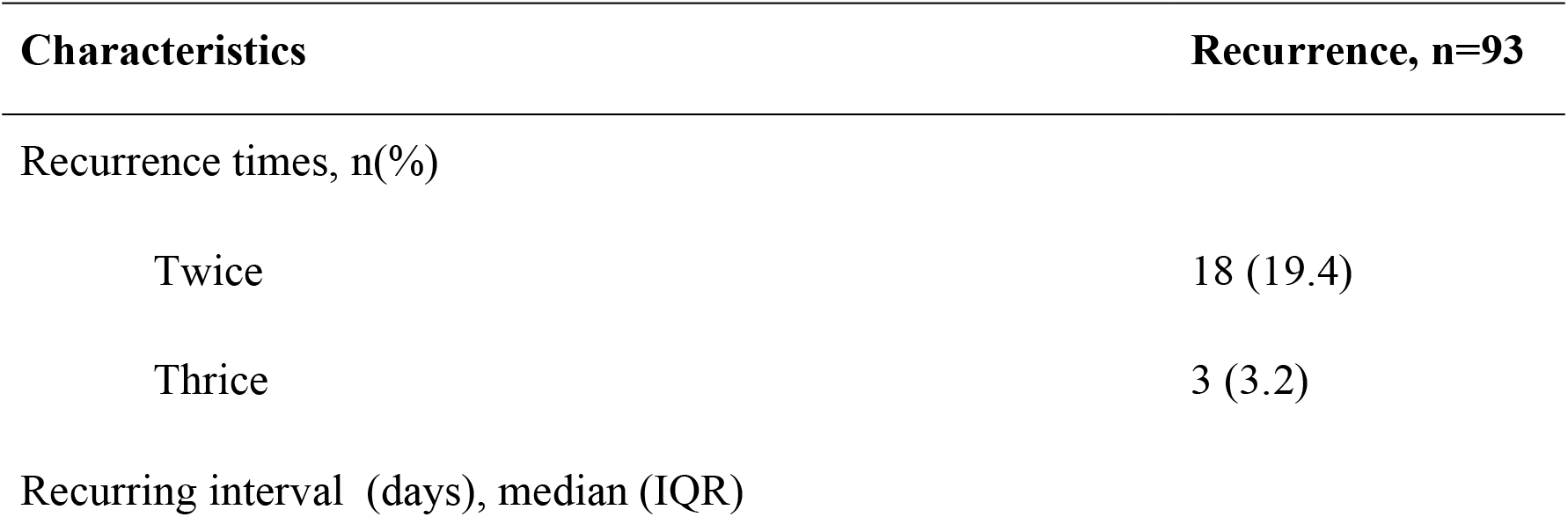
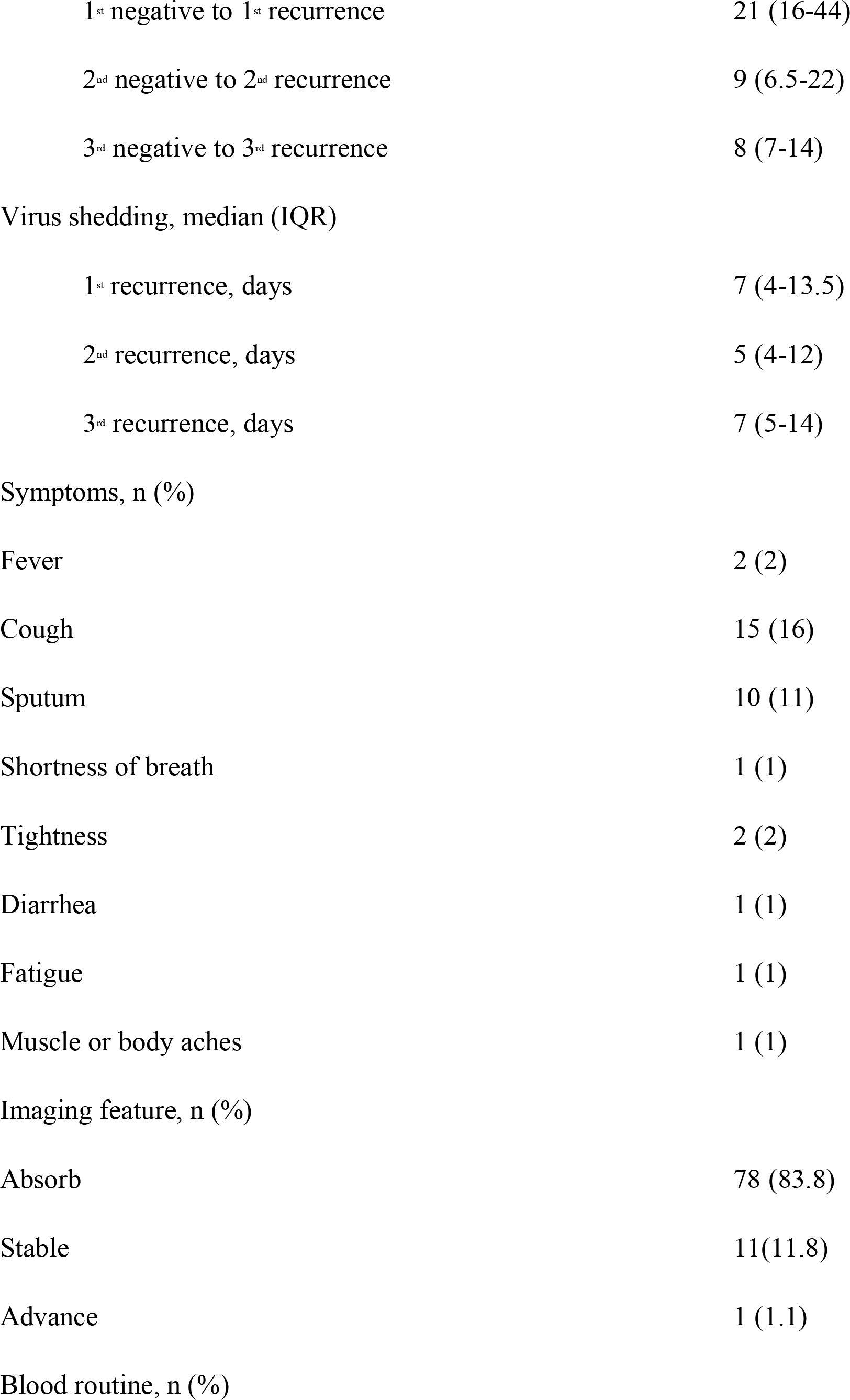
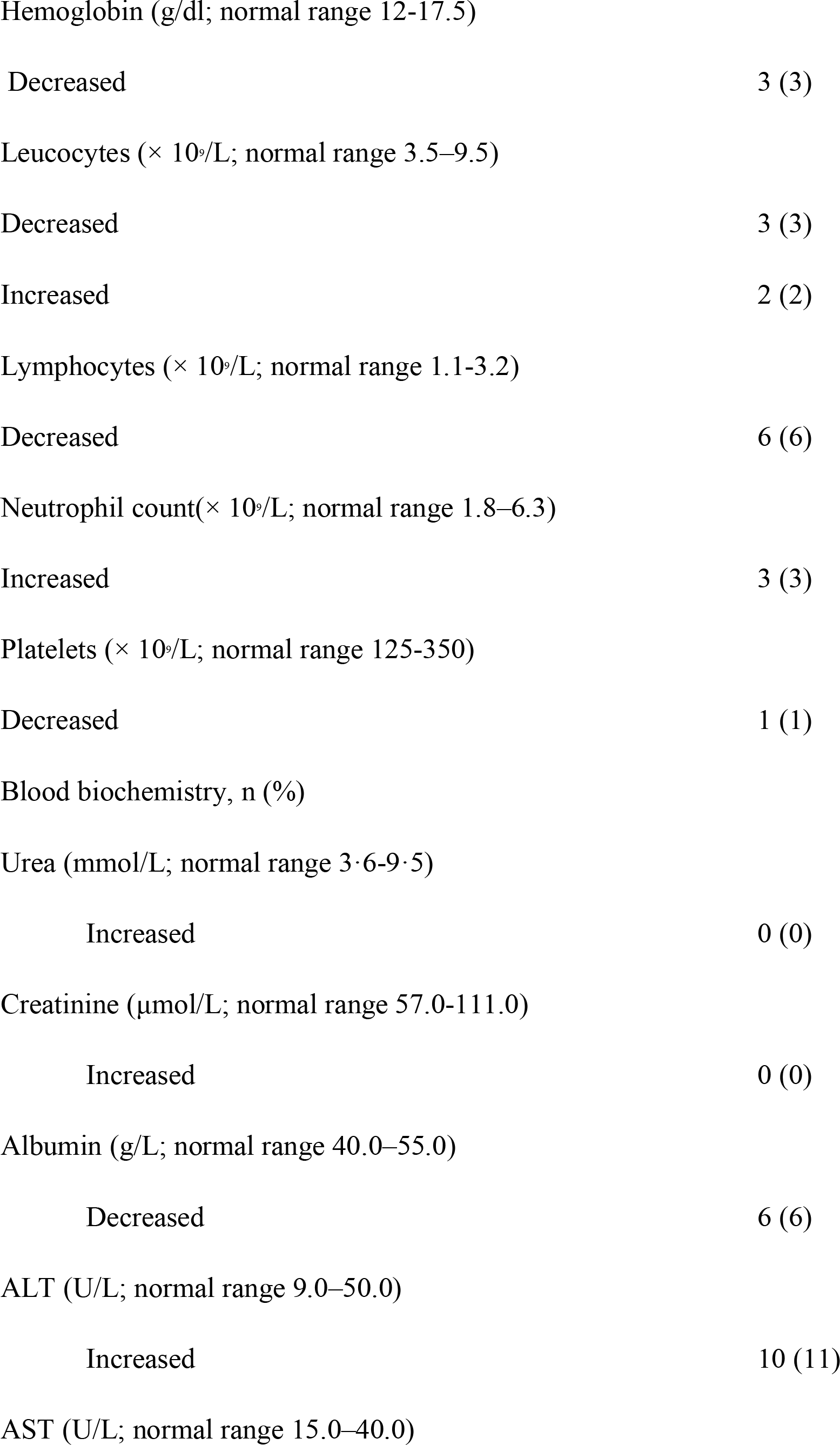
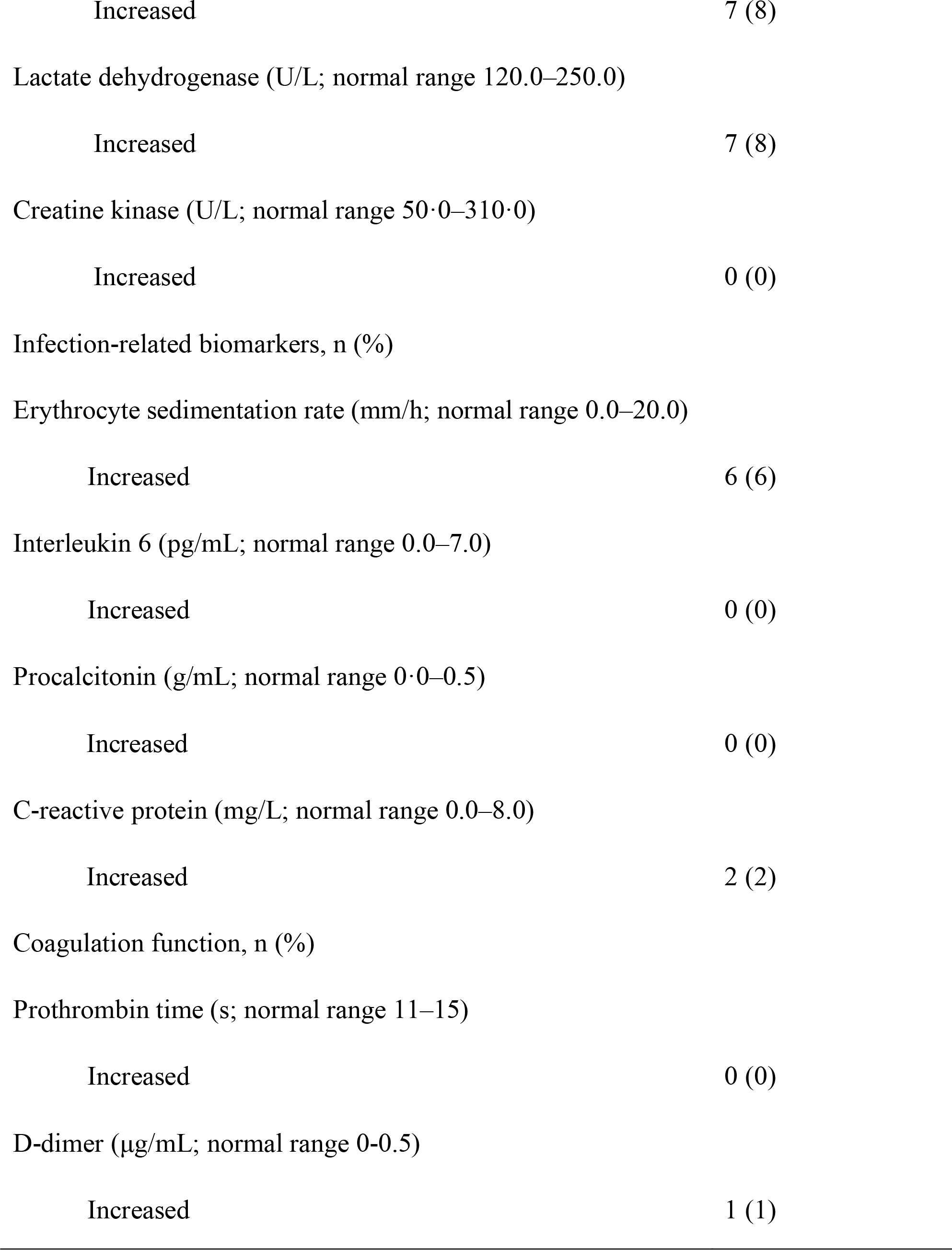
Clinical characteristics of recurring patients at follow-up. Data are n (%), n/N (%), mean (SD), and median (IQR)

**Table S3:** The effect of different drug treatments on virus lasting time

|  | **5** | **6** | **56** | **156** | **356** | **456** | **156+** | **the rest** |
| --- | --- | --- | --- | --- | --- | --- | --- | --- |
| **sample size** | 34 | 31 | 116 | 45 | 31 | 34 | 37 | 86 |
| **virus lasting time\*** | 19.85 | 24.23 | 22.72 | 29.20 | 17.61 | 25.21 | 36.73 | 25.23 |
| **second in hospital stays** | 5.29 | 4.28 | 3.34 | 0.96 | 1.68 | 2.97 | 2.54 | 3.51 |
| **sum\*\*** | 25.15 | 28.51 | 26.06 | 30.16 | 19.29 | 28.18 | 39.27 | 28.74 |

\* virus lasting time before the first discharge
\*\* sum = virus lasting time + second in hospital stays

**Table S4:** Marginal recurring probabilities in different drug treatments groups before and after SMOTE and VT

|  | 5 | 6 | 56 | 156 | 356 | 456 | 156+ | the rest |
| --- | --- | --- | --- | --- | --- | --- | --- | --- |
| original recurring prob. | 0.214 | 0.286 | 0.213 | 0.133 | 0.172 | 0.167 | 0.176 | 0.266 |
| recurring prob. after SMOTE and VT | 0.208 | 0.265 | 0.193 | 0.117 | 0.188 | 0.170 | 0.168 | 0.255 |

**Table S5:** Discovered preferred dug combinations in each age groups of normal and overweight COVID-19 patients

| **BMI** | **Age** | **Drug Preference** |
| --- | --- | --- |
| **normal weight** | 20 - 25 | 156+ |
|  | 25 - 33 | 156, 456 or 156+ |
|  | 33 - 65 | 156 |
|  | 65 - 70 | 56, 156 or 156+ |
| **overweight** | 20 - 50 | 456 |
|  | 50 - 70 | 56 |

**Table S6:**
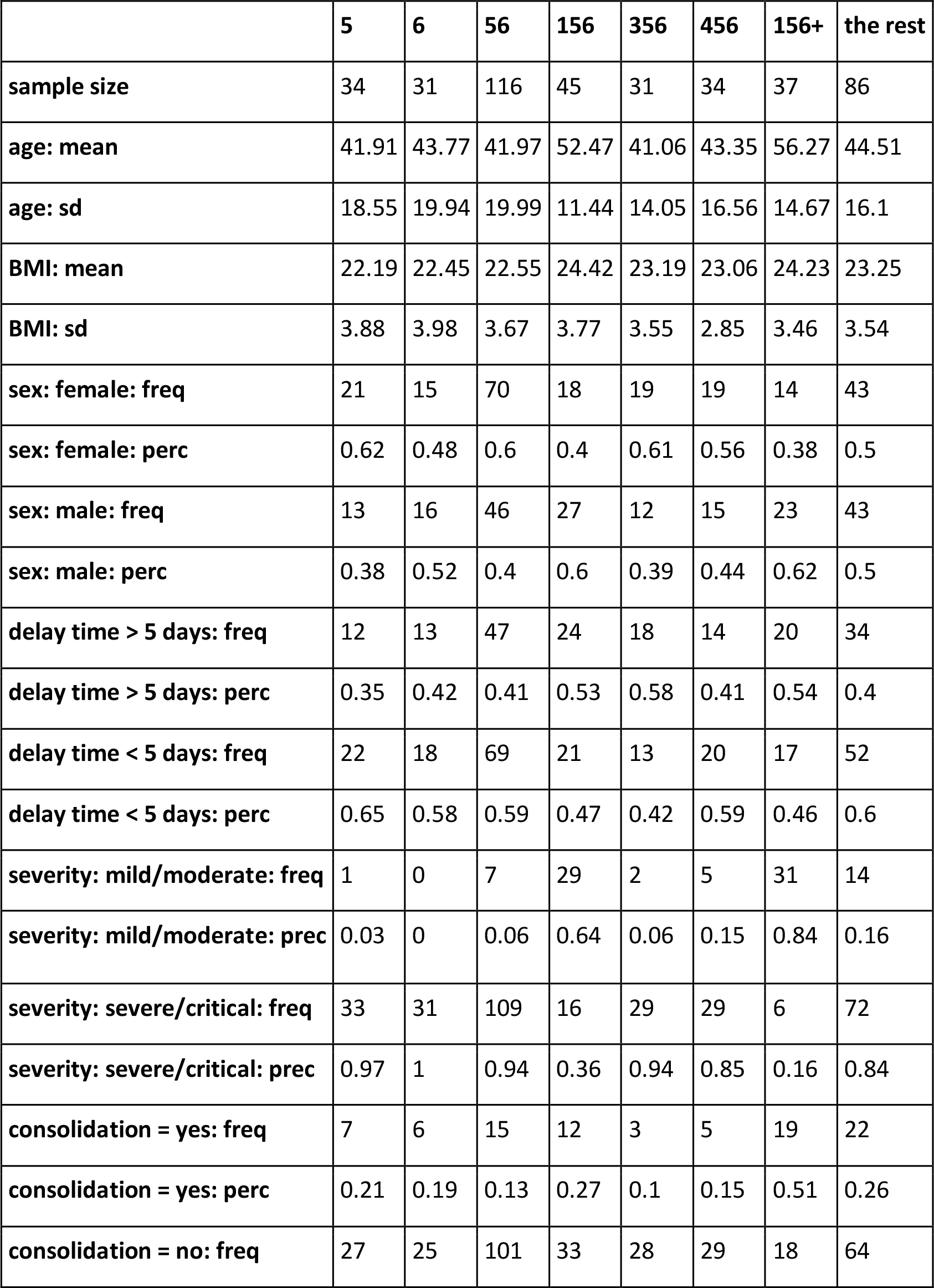
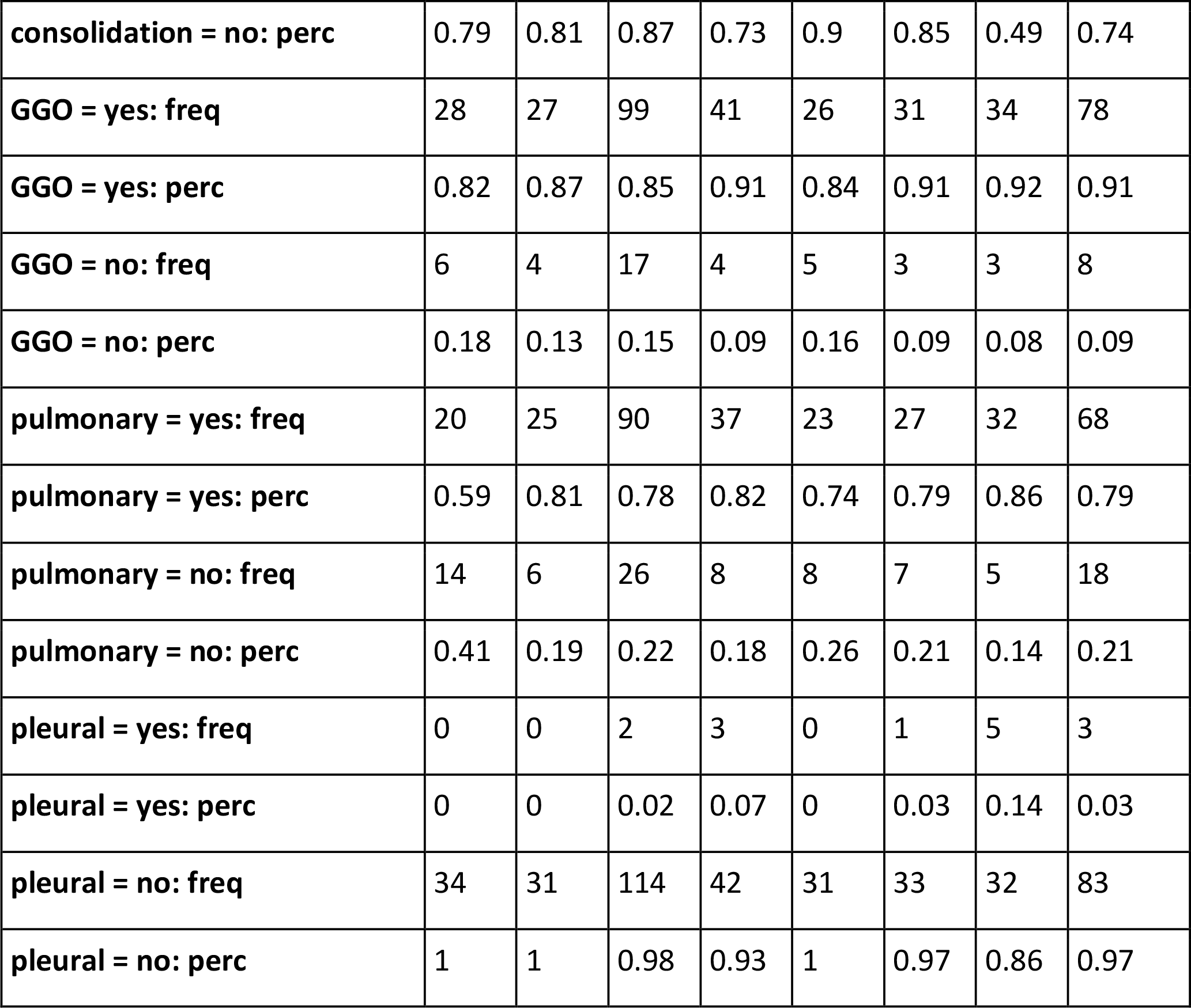
Demographics and baseline characteristics of COVID-19 patients in different drug treatments. Continuous variables (i.e., age and BMI) are shown as means with SDs, categorical variables (e.g., sex, severity…) as frequencies and percentages

